# Integrated molecular analysis of NSCLC brain metastasis tissue and multimodal ctDNA reveals distinct signatures of patient outcomes

**DOI:** 10.64898/2026.06.29.26355802

**Authors:** Darin Dolezal, Sampada Chande, Giancarlo Bonora, Yong Huang, Myles Walsh, Savannah Kandigian, Wei Wei, Anna Arnal-Estapé, Kurt A. Schalper, Sarah Goldberg, Darren Cross, Massimo Squatrito, Nicholas A. Blondin, Shidong Jia, Veronica L. Chiang, Don X. Nguyen

## Abstract

While recent therapeutic advances have extended the survival of patients with non-small cell lung cancer (NSCLC), overcoming metastatic progression in the CNS remains a significant challenge. Some patients with NSCLC may require concurrent management of CNS and extracranial metastases, while others develop isolated brain metastasis or leptomeningeal disease. These heterogenous clinical outcomes are difficult to predict and diagnose for early intervention with current surveillance modalities. Herein, we comprehensively analyzed gene mutations, copy number variations, and DNA methylation of NSCLC brain metastasis tissue collected at the time of craniotomy, combined with ctDNA sequencing of paired plasma and CSF liquid biopsies. We confirmed a high concordance between the molecular features of brain metastasis tissue with ctDNA from CSF which were largely distinct from ctDNA alterations in paired plasma samples. Plasma ctDNA tumor fraction and ctDNA hypermethylation were most significantly associated with extracranial metastasis and overall survival. Alternatively, we identified specific hypermethylated DNA loci in brain metastasis tissue and CSF ctDNA as significant correlates of brain metastasis progression and risk of leptomeningeal disease. Our findings support the utility of integrating ctDNA testing from CSF and plasma, while revealing distinct epigenetic features and biomarkers of brain metastasis or leptomeningeal disease.

**One Sentence Summary:** DNA methylation signatures in brain metastasis tissue and CSF ctDNA predicts CNS progression and leptomeningeal disease.

## INTRODUCTION

Central nervous system (CNS) metastases are present in approximately 25% of patients with non-small cell lung cancer (NSCLC) at diagnosis (*1*), and the incidence rises even further in patients who progress while on therapy (*2–5*). Historically, CNS metastasis has been associated with high morbidity, disparities in access to care, and poor survival. Yet, as new systemic therapies prolong survival, patients with CNS metastases now have a wide range of clinical outcomes, which can be difficult to anticipate and diagnose (*6*). CNS metastases are most commonly detected in the brain parenchyma (or brain parenchymal metastasis; BrM). Leptomeningeal metastasis (LM) occurs in approximately 5% of NSCLC cases (*7*), rising up to 15-20% in patients who are refractory to treatment (*8, 9*). In LM, malignant cells infiltrate the meningeal arachnoid and pia mater, as well as the subarachnoid space (SAS), which encases the brain and spinal cord and is filled with cerebrospinal fluid (CSF) (*9*). Patients with LM often have worse neurological symptoms and poorer prognosis than patients with BrM alone (*10*). LM diagnosis remains imprecise despite a rise in its incidence. Moreover, very little is understood about the underlying molecular drivers and biomarkers of BrM and LM progression and treatment options for patients with advanced LM are very limited.

Prior studies comparing single nucleotide tumor variants (SNVs) or copy number variations (CNVs) in brain metastasis to matched extra-cranial tumors found sub-clonal driver mutations to be enriched (but not necessarily specific) in BrM tissue (*11, 12*). Nevertheless, the challenges associated with safely obtaining central nervous system tissue restricts its routine use for biomarker testing. Alternatively, cell-free DNA (cfDNA) from various biofluids contains circulating tumor-derived DNA (ctDNA) and can be used to identify somatic genomic alterations (*13*). Approaches to sequencing ctDNA from plasma have emerged as noninvasive methods for monitoring minimal residual disease (MRD) as well as detecting recurrence. ctDNA from CSF is also a robust substrate for targeted or tissue-informed SNV detection and the ctDNA mutation profile of CSF better recapitulates the genomic alterations in matched CNS tumor tissue (*14–16*). More recently, tumor-agnostic approaches reveal genome-wide molecular features of ctDNA, including copy number aberration and tumor methylation patterns that can be utilized for diagnosis and classification of CNS tumors (*17–19*). Importantly, some patients with lung cancer and BrM require concurrent management of CNS and systemic disease, whereas others may develop isolated BrM or LM after initial treatment. These distinct clinical scenarios require an integrated approach to monitor tissue biopsies, CSF, and plasma for improved clinical surveillance and treatment response assessment. However, to date, few studies have directly compared the genomic or epigenomic landscape of ctDNA across paired CSF and plasma from patients with CNS metastasis.

Herein, we performed comparative and integrated analyses of different tumor tissue and ctDNA sequencing assays across matched BrM tissues and liquid biopsies from CSF and plasma and then ascertained their clinical impact in patients with CNS-metastatic NSCLC.

## RESULTS

### Genomic and epigenomic alterations in surgically resected BrM from NSCLC

Brain metastasis tissue, CSF, and plasma were collected at the time of open neurosurgical intervention in patients with NSCLC and CNS metastasis. CSF was collected either via cannulation of a lateral ventricle or from an accessible adjacent subarachnoid cistern prior to tumor resection. We assessed the association between SNVs, CNVs, and differentially methylated fragments (DMFs) with clinicopathologic features and CNS-related patient outcomes following surgery **(Fig. 1A)**. The study design included two independent cohorts (**Table 1**); cases with matched tumor tissue and CSF/plasma were first analyzed (cohort 1; 55 patients; clinicopathologic and molecular features summarized in **Tables S1-S2**) and key findings validated on a non-overlapping set of BrM tissue from craniotomy (cohort 2; 45 patients; **Table S3**). None of the patients were diagnosed with LM at the time of craniotomy.

**Fig. 1.**
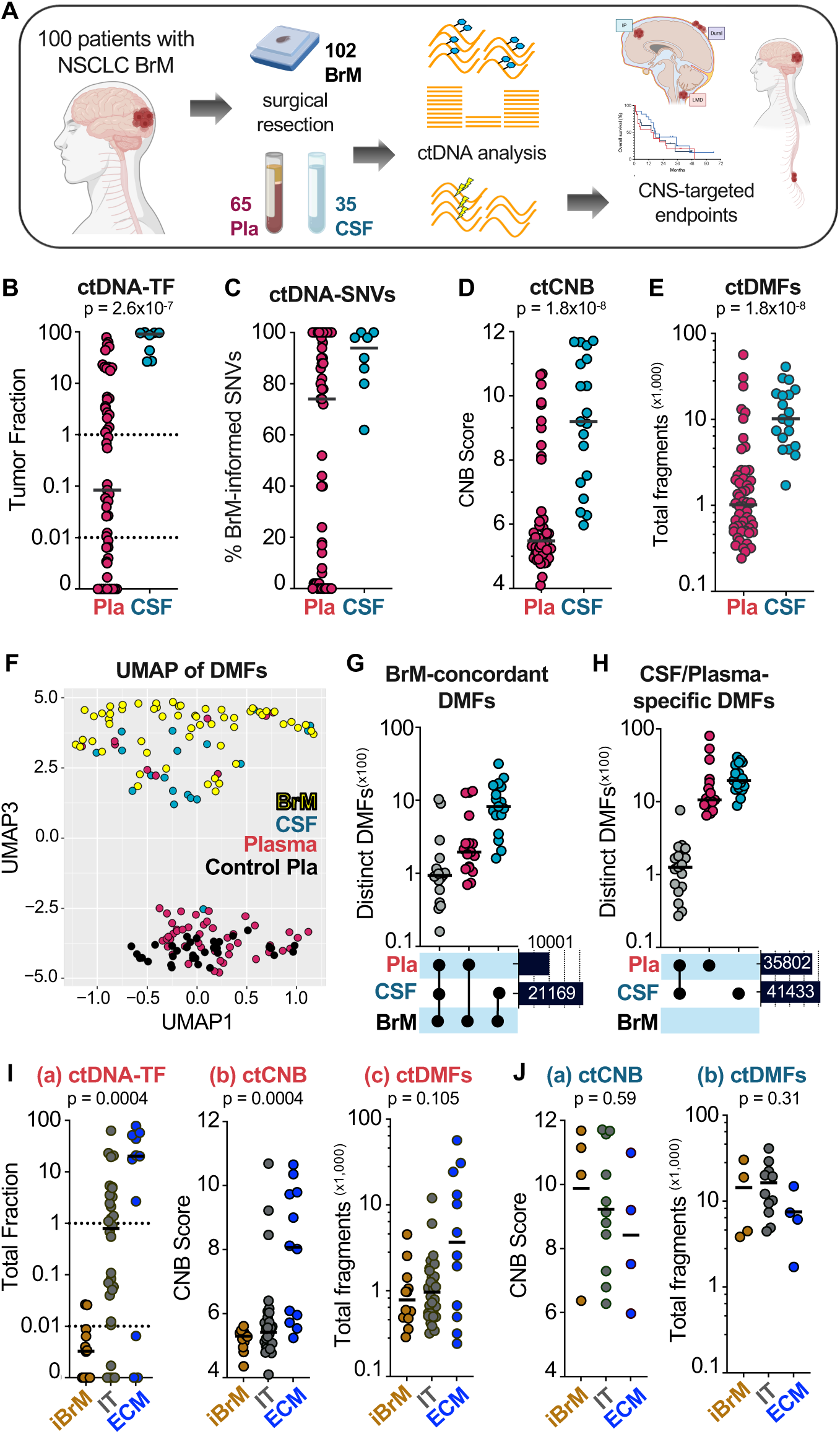
Tumor informed ctDNA alterations in cerebrospinal fluid and plasma from patients with NSCLC CNS metastasis. **(A)** Overview of study design. Abbreviations: BrM, brain parenchymal metastasis tissue. Pla, matched plasma. CSF, matched cerebral spinal fluid. ctDNA, circulating tumor DNA. **(B-C)** ctDNA tumor fractions (TF) and proportion of tumor-informed somatic variants (SNVs) detected in plasma and CSF. Data based on PredicineBEACON tumor-informed MRD assay. N=53 Pla, 8 CSF. Each dot represents an individual patient sample and bars represent median values. **(D-E)** ctDNA copy number burden (ctCNB) and differentially methylated DNA fragments (ctDMFs) were measured in plasma and CSF on PredicineEPIC methylation profiling assay. N=54 Pla, 19 CSF. P value calculated by Mann-Whitney test. **(F)** Uniform manifold approximation and projection (UMAP) of CSF, plasma, and BrM samples based on genomic locations of DMFs. Each point represents an individual sample. Samples color coded by specimen type. Control plasma samples from healthy donors included for comparison. **(G)** Upset plot showing concordance of DMFs detected in plasma and CSF with matched BrM tissue. Connected dots represent features that are exclusively shared among the indicated groups. N=18 patients with matched tissue, plasma, and CSF triplets. **(H)** Upset plot showing DMFs detected in plasma and CSF that are not shared with matched BrM tissue plotted as in (G). **(I)** Plasma ctDNA-TF (a), ctCNB (b), and ctDMFs (c) was plotted for patients with isolated brain metastasis (iBrM), BrM and isolated thoracic tumors (IT), or BrM and widespread extra-cranial metastasis (ECM). All values are represented and assessed using Kruskal-Wallis test. N=12 iBrM, 30 IT, 12 ECM. **(J)** CSF ctCNB (a) and ctDMFs (b) was plotted for patients as in (F). N=4 iBrM, 11 IT, 4 ECM.

**Table 1.** Study cohort. Clinicopathologic characteristics of the study patients in cohorts 1 and 2.

| Study Population Clinicopathologic Features |  |  |
| --- | --- | --- |
|  | Cohort 1 | Cohort 2 |
| No. of Patients | 55 | 45 |
| Age, Median (Range) | 66 (35 - 89) | 70 (53 - 85) |
| Sex F / M | 34 (62%) / 21 (38%) | 25 (56%) / 20 (44%) |
| Smoking Hx / Non-smoking Hx | 47 (85%) / 8 (15%) | 43 (96%) / 2 (4%) |
| Pack-years, Median (Range) | 35 (0 - 125) | 35 (0 - 80) |
| KPS Score, Median (Range) | 75 (60 - 100) | 70 (60 - 100) |
| Post- or On- Treatment | 34 (62%) | 24 (53%) |
| No Prior Treatment | 21 (38%) | 21 (47%) |
| **BrM tumor features** |  |  |
| Size of Resected BrM, Median (Range) | 3.5 (1.2 - 7.2) | 3.3 (1.6 - 7.8) |
| No. of BrMs: 1 | 23 (42%) | 18 (40%) |
| 2-to-3 | 11 (20%) | 14 (31%) |
| >3 | 21 (38%) | 13 (28%) |
| Posterior Fossa involved | 14 (25%) | 8 (18%) |
| Isolated BrM (iBrM) | 14 (25%) | 9 (20%) |
| Extracranial Metastasis (ECM) | 12 (22%) | 16 (36%) |
| Adenocarcinoma | 38 (69%) | 33 (73%) |
| Non-adenocarcinoma, NSCC | 17 (31%) | 14 (31%) |
| EGFR Activating Mutation | 10 (18%) | 2 (4%) |
| ALK translocation | 1 (2%) | 0 (4%) |
| **Clinical follow-up** |  |  |
| Median Follow-up Time | 39 months | 54 months |
| Median Survival Time | 16 months | 10 months |
| Patient Deaths | 44 (80%) | 37 (82%) |
| Systemic Progression | 20 (36%) | not recorded |
| CNS Disease Progression | 31 (56%) | 28 (62%) |
| Parenchymal BrM Progression | 19 (35%) | 21 (47%) |
| Leptomeningeal Progression | 12 (22%) | 7 (16%) |

Whole exome sequencing (PredicineWES+) of resected BrM was performed to identify baseline mutations for tumor-informed targeted liquid biopsy assays (Cohort 1; **Table S4**). Non-silent somatic SNVs were detected across 8,192 genes including 302 known pathogenic variants (**Fig. S1A, Table S5**). CNV analysis revealed 482 gene-level amplifications, 990 heterozygous- and 151 homozygous-deletions (**Fig. S1B, Table S6**) as well as structural variants such as ALK (1) and ROS1 (2) rearrangements (**Table S7**). The most frequent genetic alterations and affected pathways were similar to known driver mutations in primary NSCLCs (**Fig. S1C**) and were concordant with previous results from targeted sequencing of 322 NSCLC BrM craniotomies (*12*). Next, whole-genome methylation profiling (PredicineEPIC) was used to identify DMFs in BrM tissue. We detected substantial inter-tumor variation in total DMF counts per tumor, ranging from 7779 to 118,573 in cohort 1 (Interquartile range (IQR): 22817-52339) and 3196 to 110,338 in cohort 2 (IQR: 17539-50737; **Fig. S1D**). Because aggregate methylation is sensitive to chromosomal rearrangement and dosage, we assessed whether DMF counts were linked to increased chromosomal instability (Copy number burden (CNB) score). There was a weak relationship between DMF counts and CNB scores in cohort 1 (Spearman rank correlation coefficient (ρ)=0.21, 95%CI=-0.058-0.45, p=0.11) and moderate correlation in cohort 2 (ρ=0.507, 95%CI=0.24-0.70, p=0.0004, **Fig. S1E**). Altogether, this suggests large variance in methylation across NSCLC BrM and partial independence from chromosomal instability.

### Methylation and genomic alterations in CSF and plasma

Having completed a comprehensive genomic and epigenomic assessment of BrM tissue, we quantified the overall abundance of tumor-derived SNVs, CNVs, and DMFs in matching cfDNA. cfDNA was successfully extracted from 80% of CSF (28 of 35) and from all plasma samples. ctDNA-tumor fraction (TF) was estimated by applying a variant allele frequency (VAF)-based model across detectable SNVs. ctDNA-TF was significantly higher in CSF compared to plasma with median 91% versus 0.084% (p=2.6x10^-7^; **Fig. 1B**). Across tumor-informed panels, higher proportions of BrM-concordant SNVs were detected in CSF compared to plasma with median 94% versus 74% (**Fig. 1C**). ctCNB scores calculated from detectable CNVs were higher in CSF compared to plasma (p=1.8x10^-8^; **Fig. 1D**). Finally, ctDMF total fragment counts were substantially higher in CSF compared to plasma with a median of 10,199 versus 1,019 fragments (p=1.8x10^-8^; **Fig. 1E**). Uniform manifold approximation projection (UMAP) analysis based on the genomic locations of DMFs in each sample revealed close overlap of CSF and BrM tissue, whereas plasma showed more divergence and overlap with healthy control samples (**Fig. 1F**). To compare the concordance of BrM DMFs with CSF- and plasma-specific DMFs, we performed set-intersection analyses across matched tissue-CSF-plasma triplets. CSF DMFs demonstrated greater concordance with BrM tissue than plasma DMFs, with a median of 1,048 versus 197 shared fragment sites, respectively (**Fig. 1G**). Both plasma and CSF harbored DMFs not detected in BrM tissue and there was limited overlap between plasma and CSF compartments (**Fig. 1H**). These results highlight the abundance of CSF ctDNA present at time of craniotomy and its genetic/epigenetic similarity to BrM tissue.

We next compared BrM tissue and ctDNA alterations across patients with either isolated BrM (iBrM) (i.e. with previously treated and well-controlled systemic disease), BrM with intra-thoracic tumors (IT), or BrM with concurrent extra-cranial metastasis (ECM) (**Table S8**). Lower plasma ctDNA-TF, CNB, and DMFs were seen in cases with isolated BrM (iBrM), while increased levels were detected in cases with BrM and concurrent ECM (**Fig. 1I**). Conversely, CSF ctDNA-TF, CNB, and DMF counts were not associated with ECM (**Fig. J**). Thus, in patients with NSCLC and CNS metastasis, the molecular landscape of ctDNA and its relative representation of brain metastasis alterations is predicated by the source of ctDNA, extent of systemic disease, and type of molecular alteration (genetic or epigenetic).

### Plasma ctDNA methylation correlates with poor survival in NSCLC patients with BrM

BrM tissue tumor mutational burden (TMB), CNB, or DMFs did not correlate with overall survival (OS) using the median cohort values for patient stratification (**Fig. S2A–C**). Likewise, no substantial OS differences were seen across CSF ctDNA levels (**Fig. S2D–F**). Because plasma ctDNA profiling likely reflects BrM as well as systemic disease burden, we correlated plasma ctDNA features at time of surgery with patient survival. Plasma ctDNA-TF detected by tumor-informed panel testing was significantly correlated with OS (TF>0.01% hazard ratio (HR): 2.79; CI: 1.45-5.34; p=0.0020; **Fig. 2A**). We then assessed whether tumor-agnostic approaches to measure ctCNVs and ctDMFs correlated with OS. Plasma CNB did not correlate with patient survival (**Fig. S3A**), but elevated plasma ctDMFs (top two tertiles) was significantly associated with shorter survival following surgery (HR: 2.30, CI: 1.21-4.35, p=0.011, **Fig. 2B**). Elevated plasma ctDNA-TF and ctDMFs were each associated with shorter OS in univariate Cox models (**Fig. 2C, top**) and their prognostic value was independent of clinical factors in multivariate Cox models (**Fig. 2C, bottom** and **Fig. S3B**). We next investigated the relationship between plasma ctDNA alterations and subsequent CNS progression. Levels of plasma ctDNA-TF, ctCNB, and ctDMFs showed no significant association with the frequency of CNS progression events after surgery (**Fig. 2D**) or with the cumulative incidence of CNS progression over time (**Figs. 2E and S3C-D**). We conclude that the plasma ctDNA TF and methylation profile show independent prognostic value in patients with NSCLC BrM.

**Fig. 2.**
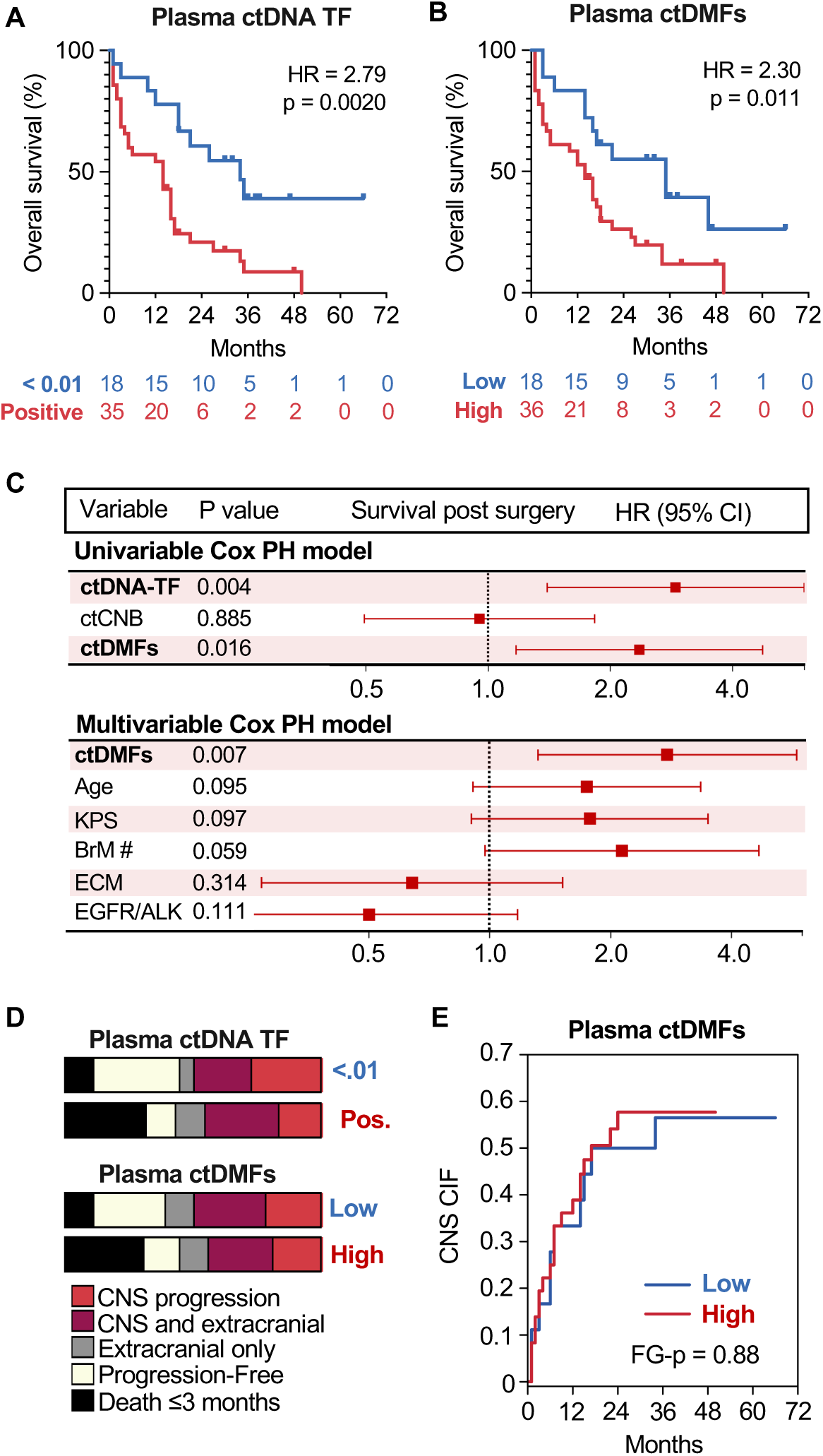
Methylation burden in plasma correlates with overall survival but not CNS progression. **(A-B)** Kaplan-Meier curves of patient overall survival (OS) by plasma ctDNA-TFs in (A) or plasma ctDMFs in (B). DMF scores were divided into tertiles to generate low-, intermediate-, and high-risk groups; the intermediate- and high-risk groups were subsequently combined into a single category. Hazard ratios (HR) were estimated using log-rank method and P values by log-rank test. CI, 95% confidence interval of HR. Numbers at risk are displayed below the x-axis. **(C)** Univariate and multivariate Cox proportional hazards regression of OS by plasma ctDNA-TF, ctCNB, and ctDMFs. All co-variables were modeled categorically: median age (>65), median KPS score (≤70), BrM# (>1), ECM (Present/Absent), and EGFR or ALK oncogenic alteration (Present/Absent). P values were calculated using a Wald test. **(D)** Stacked bar charts (100%) showing proportions of CNS and systemic disease progression events stratified by plasma ctDNA-TF and ctDMF levels. **(E)** Cumulative incidence function (CIF) of CNS progression stratified by plasma ctDMFs with death treated as a competing risk. P value calculated using a Fine-Gray (FG) model. N=36 Intermediate/High-Risk group, 18 Low-Risk group.

### Molecular markers of CNS metastasis progression

The apparent discordance between molecular correlates of overall survival and CNS progression is likely to be confounded, at least in part, by the different latency and clinical morbidity of BrM with systemic disease versus isolated BrM, as well different ctDNA mutations or DMFs that can be detected from plasma or CSF. To identify specific markers of CNS progression, we focused on comparing alterations in BrM tissue and CSF ctDNA since sampling of ctDNA from CSF is a better surrogate of BrM tissue. Overall tissue TMB and CNB did not correlate with risk of CNS progression (**Fig. 3A–B**) whereas elevated DMFs were numerically associated with increased cumulative incidence and frequency of CNS progression following surgery, but this did not reach statistically significance (> median HR: 1.69, CI: 0.85-3.35, p=0.13) (**Fig. 3C, F)**. Likewise, in CSF, ctCNB did not correlate with risk of CNS progression (**Fig. 3D**) whereas elevated ctDMFs were associated with increased cumulative incidence and frequency of CNS progression following surgery without reaching significance (> median HR: 1.45, p=0.60, **Fig. 3E–F**). Univariate Cox proportional hazards regression failed to identify any individual SNVs or CNVs in BrM tissue (from cohort 1) that were significantly associated with risk of CNS progression (**Fig. S4A-C and Tables S9–10**). Alternately, we identified 2589 specific DMFs that were significantly associated with increased risk of CNS progression (q<0.20 (p=0.013); **Fig. S4D and Table S11**). We also aggregated DMF at overlapping/adjacent loci and annotated them by their respective gene loci (gene linked DMFs ; glDMFs). This identified 359 glDMFs that were significantly associated with increased risk of CNS progression (q<0.20 (p=0.0098); **Fig. S4E and Table S12**). By contrast, few DMFs/glDMFs were associated with a decreased risk of CNS progression (**Fig. S4F–G**).

**Fig. 3.**
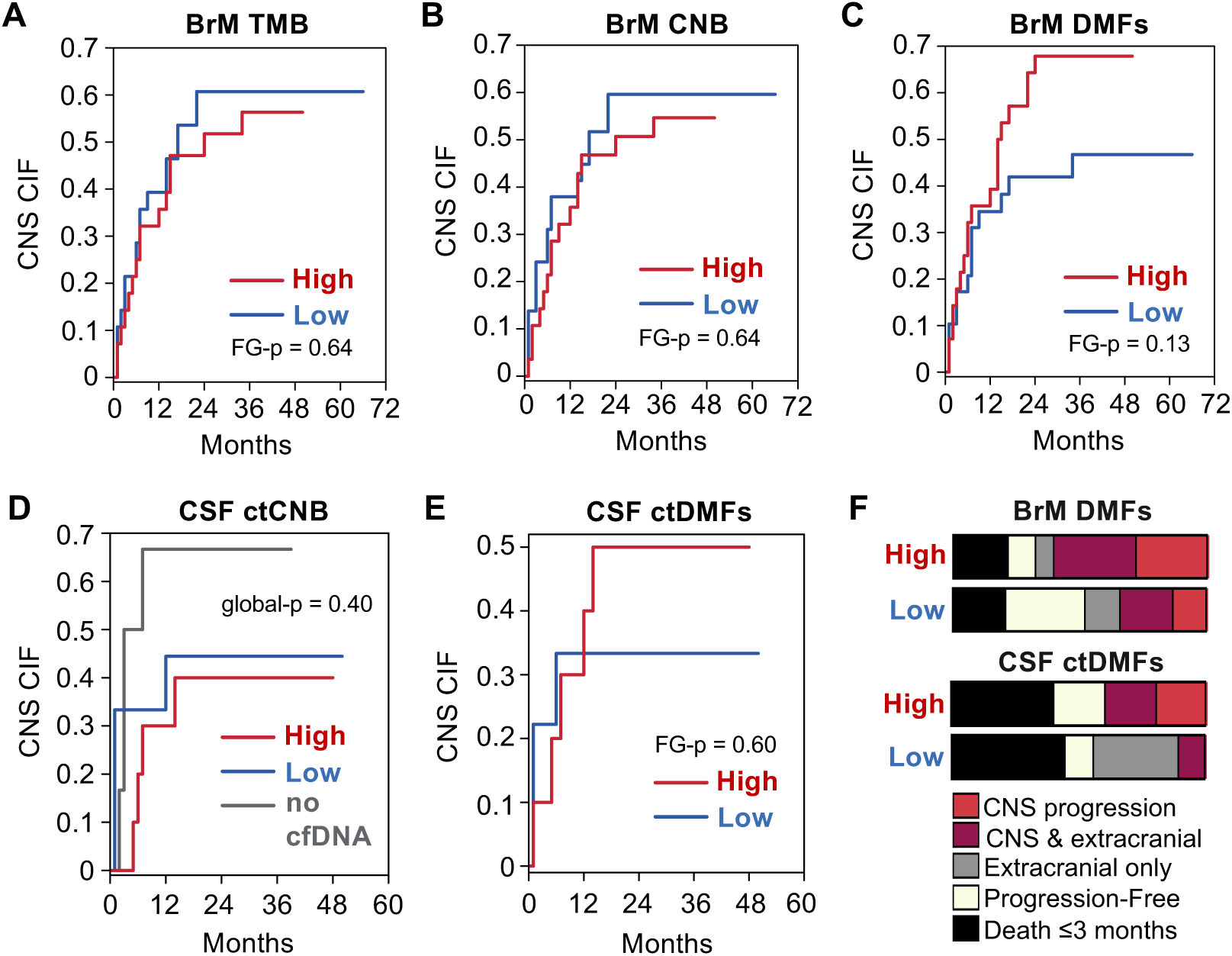
Methylation burden in tumor tissue and CSF correlates with CNS progression. **(A)** CIF of CNS progression stratified by the median BrM tumor mutation burden (TMB score). Death was treated as a competing risk. N=28 High, 28 Low. **(B)** CIF of CNS progression stratified by median BrM tissue CNB score. N=28 High, 29 Low. **(C)** CIF of CNS progression stratified by median BrM tissue DMF count. N=28 High, 29 Low. **(D)** CIF of CNS progression stratified by median CSF ctCNB score. N=10 High, 9 Low, 6 No cfDNA. **(E)** CIF of CNS progression stratified by median CSF ctDMF count. N=10 High, 9 Low. **(F)** Stacked bar charts (100%) showing proportions of CNS and systemic disease progression events by BrM tissue DMF and CSF ctDMF levels. All P values calculated by Fine-Gray model.

### DNA methylation signatures for CNS and LM progression

Because the coverage of the DNA methylome assay was broader than mutational profiling and the fact that bulk or specific DMFs were moderately correlated with CNS progression, we hypothesized that a DNA methylation signature to predict risk of CNS progression after surgery (CNS risk score) could be derived. To test this, we calculated a risk score based on the weighted aggregate of high-prevalence glDMFs in BrM tissue that were linked to CNS progression (Cox PH q<0.20, N=108 glDMFs; **Table S13**). Cases in cohort 1 with a positive risk score were associated with increased cumulative incidence of CNS progression (Fine-Gray regression, global Wald p=3.8x10^-5^, **Fig. 4A**). Importantly, a positive risk score was independently associated with an increased risk of CNS progression in cohort 2 (hazard ratio of 5.91; 95%CI: 1.06-32.78, Fine-Gray p=0.042; **Fig. 4B**). The median CNS risk scores of both cohorts were similar (9.99 vs. 13.82) and higher in CNS-progressing patients (**Fig. 4C**).

**Fig. 4.**
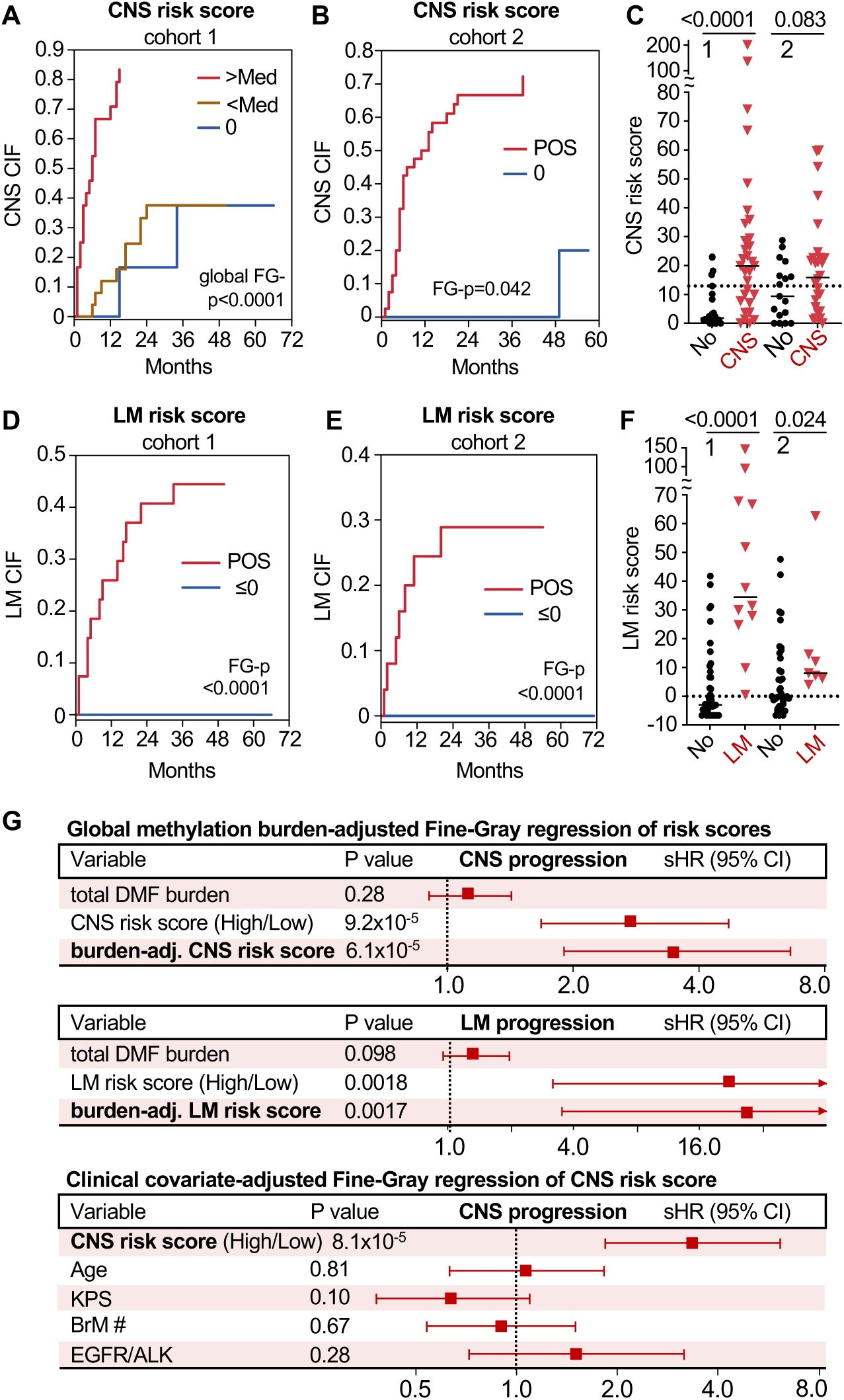
A subset of methylated DNA fragments in brain metastasis tissue is associated with CNS progression. **(A)** Gene linked DMFs that correlate with CNS progression were used to calculate a risk score (CNS risk score) from BrM tissue. CIF of CNS progression in cohort 1 patients was plotted based on their CNS risk score. Death was treated as a competing risk. P value by Fine-Gray regression calculated using omnibus Wald test for global comparison (global FG-p=3.8x10^-5^). N=55 patients (24 High^+^ / 25 Low^+^ / 6=0). **(B)** CIF of CNS progression in cohort 2 patients stratified by their BrM tissue CNS risk score (HR = 5.91; 95%CI: 1.06-32.78, Fine-Gray p=0.042). N= 45 patients (40 Positive / 5=0). **(C)** Dot plots of CNS risk scores across cohorts 1 and 2 grouped by CNS progression (red, N=59) or No CNS progression (black, N=41). Each point represents an individual sample. Bars represent group median values. P value by Mann-Whitney test. **(D)** Gene linked DMFs that correlate with LM were used to calculate a risk score (LM risk score) from BrM tissue. Cumulative incidence function (CIF) of LM specific progression in cohort 1 patients was plotted based on their LM risk score (HR=not reportable, FG-p<0.0001). N=55 patients (28 Positive / 28 ≤0). **(E)** CIF of LM progression in cohort 2 stratified by their BrM tissue LM risk score (HR=not reportable, FG-p<0.0001). N=45 patients (25 Positive / 20 ≤0). **(F)** Dot plots of LM risk scores across cohorts 1 and 2 grouped by patient by LM specific progression (red, N=19) or No LM progression (black, N=82). Each point represents an individual sample. The 0 and median values are shown as a dotted line. P value by Mann-Whitney test. **(G)** Uni- and multivariable Fine-Gray competing risks regression analyses of CNS and LM progression using CNS and LM risk scores, respectively. Global methylation burden was defined as the total DMF count per sample and modeled as a continuous Z-score. Clinical co-variables were modeled categorically: age (>65 years), KPS (≤70), BrM# (>1), and EGFR or ALK oncogenic alteration (Present/Absent). Associations with cumulative incidence of CNS or LM progression are reported as subdistribution hazard ratios (sHRs) with 95% confidence intervals, with patient death treated as a competing event. P values were calculated using Wald tests. N=100 patients (50 High / 50 Low for CNS risk score; 50 High / 50 Low for LM risk score).

In addition to progressing in the brain parenchyma, CNS metastasis can also invade into the meninges and spinal cord, leading to LM (patients with BrM-to-LM progression shown in **Table S14**). A positive CNS risk score showed a significant association with LM development in cohort 1 (Fine-Gray-p<0.0001, **Fig. S5A**) as well as in cohort 2 (Fine-Gray-p=0.0001, **Fig. S5B**), yet many cases fell into intermediate (low^+^) range. To derive a more specific predictor of LM, we calculated a LM risk score using the same approach as described above (Cox PH, p<0.01, q=0.49, N=98 glDMFs; **Table S15**). The median risk score separated patients of cohort 1 into a group with high cumulative incidence of LM (44% of cases) and a group with no LM (**Fig. 4D**). Applying this frozen threshold to the patients of cohort 2 similarly stratified groups with either high cumulative incidence of LM (28% of cases) or no LM (**Fig. 4E**). Hazard ratios could not be reliably estimated due to the complete separation of LM events. The median LM risk scores of both cohorts were similar (-0.86 vs. 2.77) and significantly higher in patients who developed LM (**Fig. 4F**). When using unannotated DMFs (i.e. not binned to gene loci; tDMFs), we found similar associations with CNS progression (173 DMFs associated with CNS progression at q<0.10; **Fig. S5C–D and Table S16**) and LM progression (334 DMFs associated with LM progression at p<0.005; **Fig. S5E–F and Table S17**) albeit with less pronounced risk stratification than glDMF signatures. Moreover, when accounting for total methylation burden in a multivariate Fine-Gray regression analysis, glDMF risk scores for CNS or LM progression remained significant (**Fig. 4G**).

tDMFs mapped to intergenic, exons, introns, 1-5 kb upstream regulatory elements, and promoter-associated regions, with the latter representing the largest proportion of methylated loci (**Fig. 5A–B**). To gain biological insight into the specificity of these alterations, we performed gene ontology analysis using the subset of glDMFs. The CNS-associated signature was enriched for genes involved in immune regulation, antiviral defense, protein homeostasis and intracellular trafficking (**Fig. 5C, Table S18**), whereas the LM-associated signature was enriched for metabolic programming, innate antiviral and interferon signaling pathways (**Fig. 5D, Table S19**). DNA methylation can have variable effects on gene transcription depending on the genomic loci. Because promoter and 5′ untranslated region methylation is most frequently linked to transcriptional repression, we performed a sub-analysis of promoter/5’UTR-linked DMFs (pDMFs). pDMFs that associated with CNS progression (Cox PH q<0.30, N=111; **Table S20**) were enriched for cellular stress response and immune-regulatory genes (**Fig. 5E, Table S21**). LM-associated pDMFs (Cox PH p<0.1, q=.52, N=91; **Table S22)** were enriched for genes involved in ion transport, cellular differentiation, and innate antiviral immune responses (**Fig. 5F, Table S23**). The consistent promoter methylation of genes involved in antiviral and interferon signaling suggests that inhibition of innate immunity may represent a common feature of CNS progression, while suppression of distinct pathways such as cellular differentiation programs may reflect biological adaptations that also facilitate LM dissemination.

**Fig. 5.**
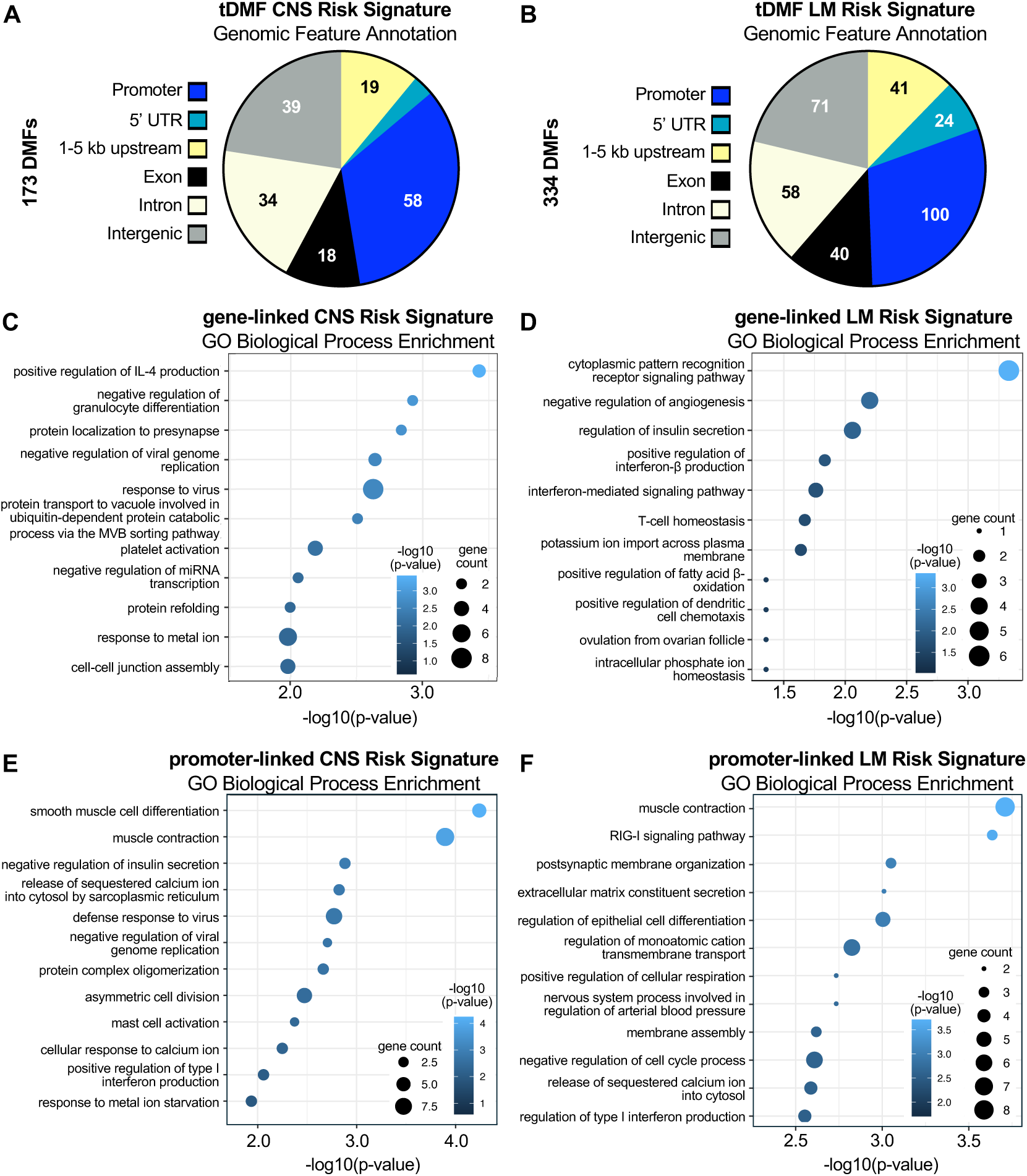
Distinct and shared biological programs underlie CNS and LM risk methylation signatures. **(A–B)** Genomic region annotations of tDMFs associated with CNS progression (A) and LM progression (B). **(C–D)** Top GO biological processes enriched within the glDMF CNS risk signature (C) and glDMF LM risk signature (D). **(E–F)** Top GO biological processes represented in the promoter-linked DMFs associated with CNS progression (E) and LM progression (F). The x-axis shows the −log10(adjusted p-value).

### CSF ctDNA methylation distinguishes LM disease from BrM

Next, we calculated risk scores of ctDMFs in CSF samples and evaluated their concordance to patient matched BrM tissue. Matched comparison of CSF:tissue pairs showed moderate but significant correlation between tissue and CSF risk scores (CNS/LM- Spearman’s ρ=0.38/ρ=0.051, p=0.053/p=0.013; **Fig. 6A**). A high CNS risk score in CSF (top two tertiles) was also associated with increased frequency and cumulative incidence of CNS progression, but this did not reach statistical significance (HR: 4.23, Fine-Gray-p: 0.20; **Fig. 6B–C**), likely due to low sampling of CSF cases. In addition to the challenges of predicting LM progression at the time of craniotomy, cytologic testing using CSF to confirm LM diagnosis is notoriously imprecise. Hence, we also analyzed ctDNA DMFs in CSF samples that were collected in patients with suspected LM and subjected to standard of care cytology. DMF counts of LM CSF were significantly higher than all BrM CSFs with a threshold of 50,000 counts able to clearly distinguish LM versus BrM CSF (**Fig. 6D**). By comparison, cytopathology examinations performed on these specimens showed 33% positive (2 of 6), 33% equivocal/suspicious (2 of 6), and 33% (2 of 6) negative for malignant cells (**Table S24**). Plasma DMF counts, however, did not significantly differ between patients who developed BrM or LM (**Fig. 6E**).

**Fig. 6.**
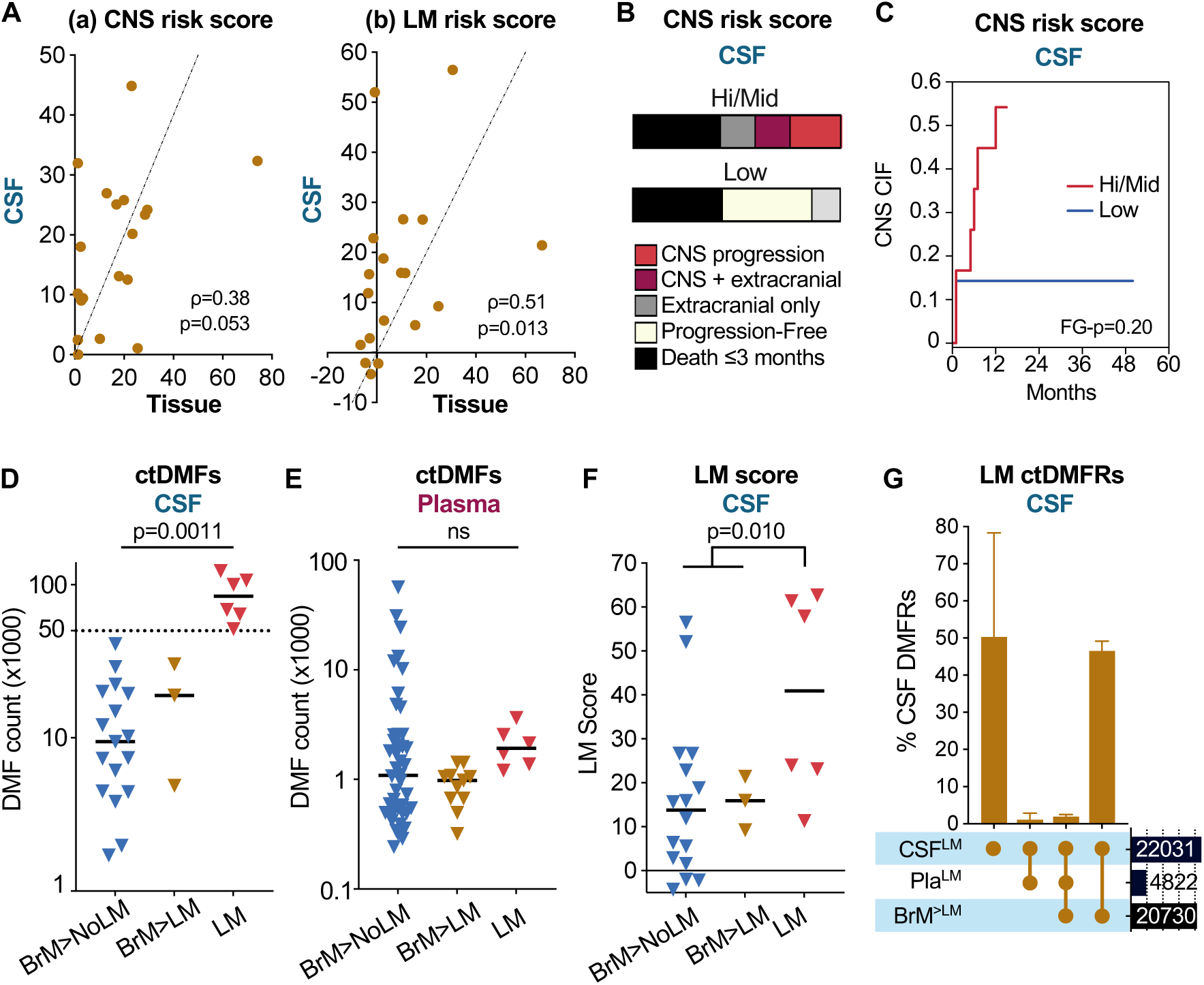
ctDNA methylation in CSF correlates with progression to leptomeningeal metastasis. **(A)** Scatter plots of CNS risk score (a) and LM risk score (b) in CSF versus matched BrM tissue. Each dot represents an individual patient’s risk score in CSF (y-axis) and tissue (x-axis). Correlation between CSF and tissue assessed by Spearman’s rank correlation coefficient (ρ) with P value. N=19 pairs. **(B)** Stacked bar charts (100%) showing proportions of CNS and systemic disease progression events stratified by CNS risk score tertiles detected in CSF. N=19 patients (12 High/Mid / 7 Low). **(C)** CIF of CNS progression in cohort 1 patients stratified by the CNS risk score detected in CSF ctDNA. Death treated as a competing risk. P value by Fine-Gray model. N=19 patients (12 High/Mid / 7 Low). **(D)** Dot plot of ctDMF counts detected in CSF collected at time of craniotomy or at LM diagnosis. BrM>NoLM: Patient with brain metastasis and no eventual LM progression, BrM>LM: Patient with brain metastasis and subsequent LM progression, LM: Patient with confirmed diagnosis of LM. Each point represents an individual sample. Bars represent median values. The dotted line represents DMF count of 50,000 fragments. N=25 total (16 BrM>NoLM, 3 BrM>LM, 6 LM). **(E)** Dot plot of ctDMF counts detected in plasma collected at craniotomy or LM diagnosis as in (D). N= 61 total (44 BrM>NoLM, 11 BrM>LM, 6 LM). **(F)** Dot plot of LM risk scores calculated from CSF collected at craniotomy or LM diagnosis as in (D). **(G)** Upset plot of ctDMFs across CSF (CSF^LM^) and plasma (Pla^LM^) collected at time of LM, compared with DMFs in matched BrM tissue from the patient’s preceding surgical resection (BrM^>LM^). N=3 patients.

LM ctDNA risk scores were further elevated in the CSF of patients with confirmed LM compared to all other groups (**Fig. 6F**). Set-intersection analysis across matched LM CSF and BrM tissue from three available cases showed approximately 50% of shared DMFs (**Fig. 6G**). To confirm if the distinct features of LM CSF are likely to represent tumor-associated alterations, we performed a sub-analysis of DMF loci that are linked to The Cancer Genome Atlas (TCGA)-associated cancer genes. This cancer enriched DMF loci analysis clustered CSF samples based on whether they were derived from LM or BrM cases (**Fig. S6A–B**). Thus, distinct BrM and LM DNA methylation profiles from CSF can potentially be used for diagnostic as well as prognostic purposes.

### Longitudinal CSF monitoring of CNS progression

As a proof of concept, we compared ctDNA readouts across various cases where longitudinal sampling of CSF and/or plasma was available. Patient 23 underwent craniotomy followed by stereotactic radiosurgery for relapsed lung adenocarcinoma brain metastasis four years after prior lobectomy and chemoradiation. BrM tissue showed high scores for CNS and LM progression. Seven months after surgery new parenchymal outgrowth was observed, followed at 14 months by MRI imaging and CSF cytology findings suspicious for LM (**Fig. 7A**). They showed no evidence of systemic disease progression after craniotomy. Although an Ommaya reservoir was placed and intrathecal chemotherapy was started, the patient passed away three months afterwards. Intraoperative ventricular CSF and Ommaya CSF samples collected before and after chemotherapy contained 8.8, 4.5, and 1.0 ng/mL of cfDNA, respectively (**Fig. 7B**). Despite having relatively lower total cfDNA, CSF collected at time of LM diagnosis contained higher fractions of total ctDNA (**Fig. 7C**). All CSF samples contained elevated CNS and LM risk scores which tracked with worsening of disease (**Fig. 7D**). CSF ctDMF counts were relatively low at craniotomy but increased at LM, both before and after the start of treatment (**Fig. 7E**). Although abundant cfDNA was present in matched plasma, no tumor variants were detected while the risk scores remained low and ctDMFs were minimal, suggesting that plasma ctDNA was not correlating with CNS progression (**Fig. 7B–E**). The patient’s CSF cytology switched from ‘suspicious’ to ‘negative’ following the start of intrathecal chemotherapy yet their CSF risk scores and total DMF counts remained high, consistent with persistent disease. In another case (Patient 101) with LM in the absence of parenchymal BrM, CSF ctDMFs and risk scores also remained elevated over 2 months (**Fig. S7A-D**). Swimmer plots show detectable CSF ctDNA at time of parenchymal BrM and subsequent LM diagnosis for many other cases (**Fig. S7E**). Altogether, our findings indicate that ctDNA methylation can be measured from CSF to track disease progression after craniotomy.

**Fig. 7.**
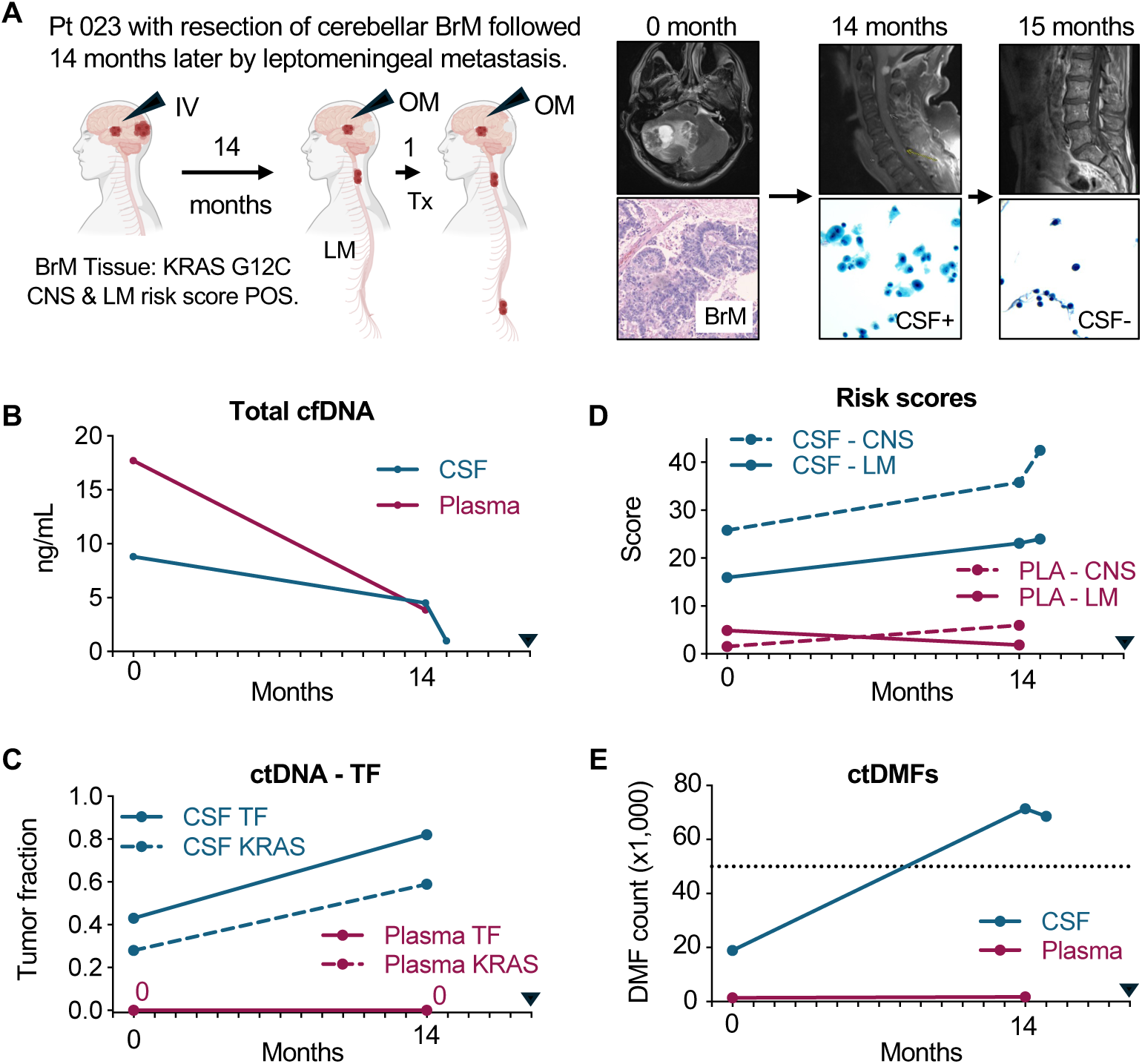
ctDNA features of CNS progression across longitudinal plasma and CSF biopsies. **(A)** Overview of clinical course, pathologic findings, and liquid biopsy sample collections from patient 34. Black arrows represent collections. IV, Intraventricular. OM, Ommaya port. Tx, Treatment. N=1 BrM tissue, 2 PLA, and 3 CSFs samples. Depicted are patient 34’s annotated H&E images of BrM tissue, magnetic resonance images of brain and spine, and cytology from lumbar puncture. **(B)** Total cell-free (cf)DNA extracted from matched CSF and plasma samples of patient 34 at time of craniotomy (t = 0 months), at diagnosis of LM (t = 14 months), and after the start of CNS-related treatment (t = 15 months). Black arrow represents time of the patient’s death. **(C)** ctDNA-TF and KRAS G12C variant allele fraction (VAF) detected across CSF and plasma sample, plotted as in B. **(D)** CNS and LM risk scores detected across CSF and plasma, plotted as in B. **(E)** Total ctDMFs detected across CSF and plasma plotted as in B. The dotted line represents DMF count of 50,000 fragments.

## DISCUSSION

We characterized genomic and epigenomic features of NSCLC BrM tumors alongside patient matched ctDNA from CSF and plasma collected at time of surgery and then followed patients longitudinally to assess for the development of additional BrM or LM. Because of this matched study design, we were able to establish the molecular concordance across these biospecimens as well as identify molecular features related to patient co-variables and outcomes post-surgery. Specifically, plasma ctDNA-TF SNVs correlated with extracranial metastasis and overall survival. Conversely, CSF ctDNA did not correlate with OS in our cohort. In a separate study examining mostly non-smoking patients and patients with prior tyrosine kinase inhibitor treatment, a significant association between CSF ctDNA and OS was reported (*20*). By contrast, our cohorts included patients with a history of smoking, CNS disease that warranted craniotomy, and who received various treatments. Therefore, overall survival based on ctDNA levels may depend on patient subgroup and their treatment history.

In addition to genetic alterations, epigenomic and discrete epigenetic alterations can drive metastasis (*21, 22*). Certain primary lung adenocarcinomas display a genome wide DNA hypermethylated phenotype (*23–25*), and DNA methylation signatures in primary NSCLC tumors can correlate with the risk of BrM development (*26*). We analyzed BrM tissues sampled directly at the time of craniotomy and identified a subset of poor prognosis BrMs that displayed increased DNA methylation. Although DNA methylation can buffer against overexpression of co-amplified adjacent genes (*25*), the DNA methylome in BrM tissue from our cohort did not correlate with chromosomal instability. Interestingly, we also identified DMFs in plasma and CSF that did not overlap with the DNA methylome of tumor tissue. It is possible that these DMFs reflect ctDNA contributions from non-malignant sources. We did not observe any strong association between particular SNVs or CNVs with CNS progression. Alternatively, increased methylation at specific genomic loci, some encoding for biologically relevant genes, correlated with CNS progression and the development of LM. The increase in DNA methylation occurred at gene promoters but also within other regulatory elements such as introns and exons. Hence, methylation at these loci is likely to affect gene expression in a variety of ways, including transcriptional activation or repression, as well as splicing, and chromatin remodeling (*27, 28*).

The utility of measuring tumor-informed plasma ctDNA at ultrasensitive thresholds in early-stage NSCLC patients has been well documented across the TRACERx (*29, 30*), LEMA (Lung cancer Early Molecular Assessment trial, NCT02894853), and LUCID (LUCID; LUng Cancer CIrculating Tumour DNA study, NCT04153526) studies (*31*). Specifically, these studies conclude that ctDNA collected pre- and/or post-operatively in Stage 0-III patients can be used to predict overall survival and risk of disease recurrence. Analogously, we show that tumor-informed ultrasensitive measurement of plasma ctDNA (PredicineBEACON) could determine prognosis of patients with BrM. Furthermore, we found that quantifying plasma ctDMFs using a tumor-agnostic approach (PredicineEPIC) can also make similar distinctions, obviating the technical challenges of designing tissue/patient bespoke ctDNA assays. However, none of the plasma biomarkers in our study reliably capture risk of CNS disease progression including in patients with isolated BrM. Instead, we identified DMFs signatures in CSF that correlate with CNS and LM progression. We speculate that the circulation of ctDNA from BrM tissue is mainly confined to the CNS. Therefore, while plasma ctDNA offers non-invasive risk stratification, it is likely that plasma ctDNA monitoring alone will not reliably track with CNS disease.

While the median overall survival of patients with BrM continues to improve (*6, 32*), the risk of developing LM also appears to be increasing. Recent reports suggest that up to one third of patients with newly diagnosed BrM may subsequently progress to LM (*33–37*). Early diagnosis and intervention for LM may significantly improve clinical outcomes (*38*), especially with recent advances in targeted therapies (e.g. antibody drug conjugates) and radiotherapies (e.g. proton therapy) (*39, 40*). Yet the diagnosis of LM continues to be fraught with the relatively poor sensitivity of MRI imaging and CSF cytology methods, as well as non-specific clinical symptoms. Assessment of CSF ctDNA by different methodologies suggest that tumor fractions are generally elevated, and greater than that of plasma (*41–43*). However, elevated CSF tumor fraction can also reflect parenchymal disease (*14, 44, 45*), which makes this metric alone insufficient to distinguish LM from BrM. Although patients with *EGFR* mutant NSCLC frequently develop leptomeningeal metastasis (*12, 46*) our cohorts included cases with a variety of driver oncogenes and the frequency of LM in our study was still relatively high (19/100, 19%), suggesting other molecular risk factors for LM progression. Based on our findings, we propose that monitoring of DNA methylation signatures from the CSF may be more sensitive than the current benchmarks for LM diagnosis and provide significant biological insight into the broader pathogenesis of LM.

This study has several limitations. Because our cohort is focused on patients who underwent craniotomy (often with symptomatic brain metastasis), our findings may not be generalizable to all patients including those with asymptomatic CNS metastases. Furthermore, we designed the study to compare liquid biopsy with matched tissue resection, and therefore cases with tumor necrosis, inflammation, or insufficient viable tumor cellularity for sequencing were excluded. This pre-analytics step may have skewed our cohort towards cases with more aggressive tumor biology or increased therapeutic resistance. Similar to prior reports (*44, 47*), we also found that a proportion of CSF samples failed to yield a sufficient amount of cfDNA for NGS testing, which highlights a technical limitation of CSF ctDNA-based approaches. Little is understood about the biological causes for fluctuating cfDNA levels in the CSF, which may influence how ctDNA fraction at any particular timepoint is interpreted. For our signature-based assessments, we reduced the probability of overfitting by limiting model complexity and cross-validated our findings in an independent cohort. Nevertheless, the number of matched CSF samples was limited. Moreover, treatments across patients were different and under standard of care, potentially introducing confounders but at the same time better reflecting real word clinical scenarios. Improved sampling of CSF and larger prospective studies could help validate the predictive value of BrM and LM risk signatures.

In conclusion, we demonstrate that for NSCLC BrM patients undergoing resection, integrating the analysis of ctDNA from plasma with CSF has significant prognostic and predictive potential, and can be tailored to different clinical scenarios. In particular, we propose that methylation signatures from BrM tissue and CSF may predict the risk of CNS progression and improve the diagnosis of LM thereby altering therapeutic strategies for successful long-term management of patients with CNS disease.

## MATERIALS AND METHODS

### Patient population

Patients undergoing therapeutic craniotomy for suspected brain metastasis between 2018-2023 at Yale New Haven Hospital (YNHH) were recruited for tissue, CSF, and blood biospecimen sample collection. Participants were required to have histologically confirmed lung adenocarcinoma or NSCLC BrM and a resected BrM tumor sample with adequate tumor cellularity to proceed with next-generation sequencing. Exclusion criteria included tissue samples with low volume, extensive cautery artifact, or abundant necrosis, as well as patients in whom a primary lung tumor origin could not be clearly established. Patient cohort 1 included cases with adequate tissue sample as well as sufficient CSF and/or plasma volume collected (from procedures spanning July 2018 - August 2023). Patient cohort 2 consisted of cases with adequate tumor tissue available for analyses (from procedures spanning November 2015 - February 2023). Specimen use in this study was approved by the Institutional Review Board of YNHH (Protocol: 2000023741). Comprehensive clinical information was collected for all patients including demographic, radiopathologic, and treatment information.

### Tissue, CSF, and plasma sample collection

Brain metastasis tumor tissue was collected at surgical resection in RPMI medium and immediately fixed in 10% neutral buffered formalin prior to paraffin embedding (FFPE), according to standard protocols. In select cases, archived FFPE tissue specimens were retrieved from the YHNN Department of Pathology archives. Hematoxylin and eosin-stained sections were reviewed by a board-certified pathologist to assess adequacy and estimate tumor cellularity. CSF samples were collected at time of surgery either via cannulation of a lateral ventricle or from an accessible adjacent subarachnoid cistern prior to tumor resection. Additional longitudinal CSF samples were collected via Ommaya reservoir or lumbar puncture. All CSF samples were centrifuged immediately after collection at 1,000 x g for 15 minutes and the supernatants aliquoted and stored at -80°C. Peripheral blood samples were collected in EDTA tubes, centrifuged at 1,000 x g for 15 minutes to extract plasma and buffy coat fraction, aliquoted and stored at -80°C.

### CNS-specific clinical outcomes

CNS progression was assessed in routine clinical care using a combination of clinical and radiopathologic assessments. Patients underwent routine surveillance with contrast-enhanced MRI of the brain, as well as orthogonal imaging modalities, including PET, when clinically indicated. Radiographic assessment of CNS progression was determined by a board-certified neuroradiologist. CNS progression was defined as the (i) local progression at the resection cavity, excluding findings attributed to treatment effect or radionecrosis, (ii) a new intraparenchymal lesion, (iii) unequivocal growth of existing intraparenchymal lesions, or (iv) LM development with clinical symptoms and suspicious MRI findings and/or positive CSF cytology findings. CNS progression events were substantiated by documented changes in clinical management. Time-to-CNS progression was the difference between the date of craniotomy and the earliest date of radiographic (or cytologic) progression. The median clinical follow-up time after craniotomy was 39 months and 54 months for cohorts 1 and 2, respectively.

### Circulating Tumor DNA Analysis and Minimal Residual Disease (MRD) Assessment

Plasma (and CSF) circulating tumor DNA (ctDNA) analysis was performed using the PredicineBEACON™ assay (Predicine, Inc., Hayward, CA), a personalized, tumor-informed minimal residual disease (MRD) liquid biopsy assay performed in a CLIA-certified and CAP-accredited laboratory. The methods have been described in Choi et al.(*48*).

The assay workflow utilizes a two-step approach. First, baseline genomic profiling is performed on tumor tissue or baseline plasma using PredicineWES+, a boosted whole-exome sequencing (WES) assay (covering 20,000 genes with boosted coverage of 600 cancer-related genes) to identify patient-specific somatic alterations. Based on the baseline profile, a personalized MRD panel is designed for each patient, targeting up to 50 identified somatic mutations. This personalized panel is combined with a fixed panel covering approximately 500 actionable and hotspot mutations to monitor for acquired resistance and potential second primary tumors. A ctDNA tumor fraction cutoff of 0.01% was used for detection, reflecting the analytical sensitivity of the assay. Complete lists of somatic variants, copy number alterations, and fusions detected in BrM tissues are provided in Supplemental Tables S4-S7.

Longitudinal plasma samples undergo ultra-deep sequencing using the customized panel, targeting a depth of 100,000× to ensure high sensitivity for low-frequency variants.

### PredicineEPIC whole-genome methylation analysis

Whole-genome DNA methylation profiles were generated by the PredicineEPIC ^TM^ assay with cfDNA input amount as low as 1ng. For library construction and methylation treatment, 5ng of material was processed with proprietary PredicineEPIC^TM^ reagents. The PredicineEPIC libraries were subjected to whole-genome sequencing at 25x coverage using paired-end 2x150bp sequencing. Paired-end sequences were first aligned to the GRCh37 reference assembly, and DNA fragments were then built by combining reads from the same molecules based on mapping locations and UMIs.

Each PredicineEPIC sample was then tested against a background model generated using samples from healthy donors in order to identify CpG sites covered by differentially methylated DNA fragments (DMFs). A set of hold-out healthy donor samples (not used in the generation of the background model) were tested in the same way to serve as negative controls. DMF scores were stratified into tertiles to generate low-, intermediate-, and high-risk groups, which were collapsed into a low- and intermediate/high-risk groups as indicated.

Full lists of differentially methylated fragments detected in tissue, CSF, and plasma samples with annotations have been deposited in the NCBI Gene Expression Omnibus (accession GSE327701).

### Copy number burden analysis

A whole-genome fragment coverage profile was generated for each PredicineEPIC sample, from which copy number burden (CNB) score was calculated. Briefly, copy number variants (CNVs) were first segmented into 1-Mb bins and normalized for GC content and mappability, followed by additional normalization using the average segment-level CNV profile derived from normal plasma samples. Arm-level CNV deviation was then quantified as the mean of normalized segment CNVs across each chromosome arm. The CNB score was calculated as the sum of the absolute z-scores of arm-level CNV deviations, followed by log2 transformation (*49*).

### CNS and LM risk score development

DMFs detected in cohort 1 were first filtered to include only those with ≥5 fragments and aggregated together based on their HUGO gene nomenclature. The resulting ‘gene-linked-DMFs’ (glDMFs) were filtered to retain only those present in ≥3 cases (high-prevalence glDMFs) and analyzed using univariate Cox proportional hazards regression analyses to assess for associations with CNS or LM progression (performed using R version 4.5.1). glDMFs most associated with CNS (FDR q<0.20) or LM (p<0.01, FDR=.49) in cohort 1 were used to build a weighted risk score by summing the regression coefficients (*β*) of glDMFs detected in each patient as follows:

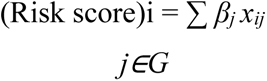

where *x_ij_* = the value of DMF *j* for patient i (defined as either 0 or 1), *β_j_* = ln(HR*_j_*) from the univariate Cox model, and *G* = set of DMFs. Cohort 1 patients were then stratified into risk groups based on the aggregate risk score. The weighted risk score and group parameters were then applied without modification to the independent patient cohort 2. Weighted risk scores were also applied to DMFs detected (at ≥2 fragment counts) in CSF and plasma. Performance was evaluated by measuring the cumulative incidence function of CNS/LM progression while treating death as a competing event. Differences between risk groups were evaluated by Fine-Gray competing risk regression with report of corresponding subdistribution hazard ratios (HR) and Wald p-values. R scripts used for Cox proportional hazards regression and risk score analyses are available from the corresponding author upon request.

### Statistical analysis

Descriptive statistics are shown as median, range, and interquartile range (IQR) as indicated. Group comparisons were made using Mann-Whitney or Kruskal-Wallis tests and two-tailed p-values reported. Spearman rank correlation was used to assess associations with DMF counts as indicated, with reporting of correlation coefficient (ρ), 95% confidence interval, and two-tailed p-value. Kaplan-Meier survival curves were compared using the log-rank test. Hazard ratios and 95% confidence intervals were estimated using Cox proportional hazards regression. Cumulative incidence function curves were compared using Fine-Gray competing-risk regression, with subdistribution hazard ratios (HR) and corresponding Wald p-values reported.

## Data Availability

Raw sequencing data generated using the PredicineWES+, PredicineBEACON, and PredicineEPIC platforms are available through the NCBI Sequence Read Archive under BioProject PRJNA1435124*. Processed PredicineEPIC data, including annotation files and matrices, and have been deposited in the NCBI Gene Expression Omnibus (accession GSE327701). All analysis code used in this study is available upon request from the corresponding author.

## List of Supplementary Materials

Fig. S1 to S7

Tables S1 to S18

Source Data

## Funding

This work was funded by the National Cancer Institute (NCI) including:

R01CA166376 (DXN)

P50CA196530 (DXN)

K12CA215110 (DD)

## Author contributions

Conceptualization: DD, VC, DXN

Methodology: GB, YH, DD, SJ

Investigation: DD, SC, GB, YH, SK

Visualization: DD, GB, YH, SK

Data curation: AA, SC, VC

Resources: NB, VC

Funding acquisition: DXN

Project administration: MW, DC, MS, SJ, DXN

Supervision: DXN, NB, SG, VC

Writing – original draft: DD, DXN

Writing – review & editing: DD, DXN

## Definitions of all symbols, abbreviations, and acronyms

CNS: central nervous system
NSCLC: non-small cell lung cancer
BrM: brain parenchymal metastasis
LM: leptomeningeal metastasis
SAS: subarachnoid space
CSF: cerebrospinal fluid
SNV: somatic nucleotide tumor variant
CNV: copy number variation
cfDNA: cell-free DNA
ctDNA: circulating tumor DNA
MRD: minimal residual disease
DMF: differentially methylated fragment
WES: whole exome sequencing
IQR: interquartile range
CNB: copy number burden score
TF: tumor fraction
VAF: variant allele fraction
UMAP: uniform manifold approximation projection
iBrM: isolated brain metastasis
IT: intra-thoracic tumor
ECM: extra-cranial metastasis
TMB: tumor mutational burden score
OS: overall survival
HR: hazard ratio
CI: 95% confidence interval
glDMF: gene-linked differentially methylated fragment
TCGA: The Cancer Genome Atlas
MRI: magnetic resonance imaging

---

**Fig. S1.**
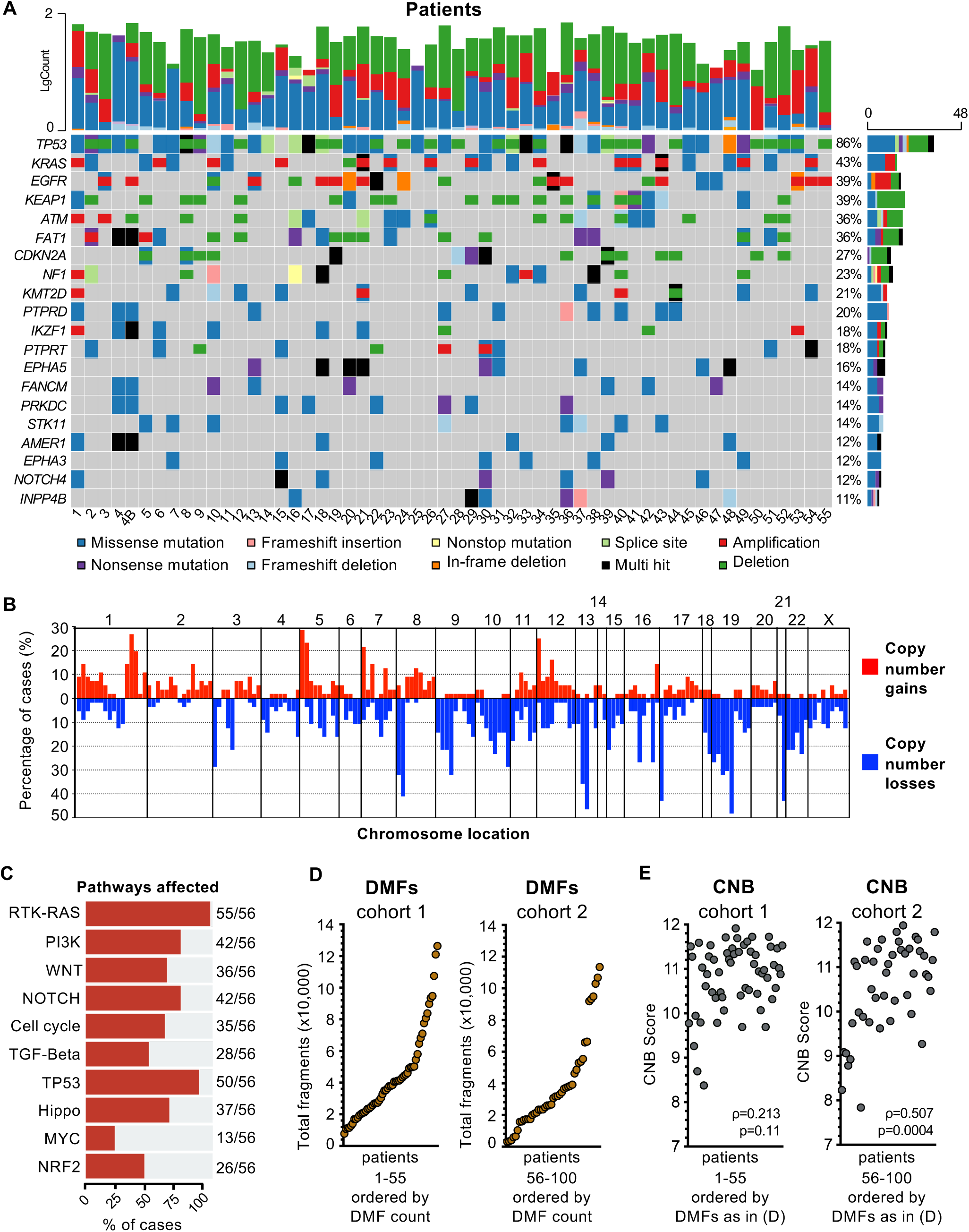
Brain metastasis tissue molecular alterations. **(A)** Oncoprint displaying the most common pathogenic alterations detected in brain parenchymal metastasis (BrM) tissue samples from cohort 1. N=56 craniotomy cases. **(B)** Copy number gains and losses detected in BrM plotted by chromosome location. **(C)** The oncogenic signaling pathways most frequently observed in BrM samples. **(D-E)** Total DMFs and CNB measured in tissue on PredicineEPIC methylation profiling assay. Each point represents an individual sample from either cohort 1 or 2.

**Fig. S2.**
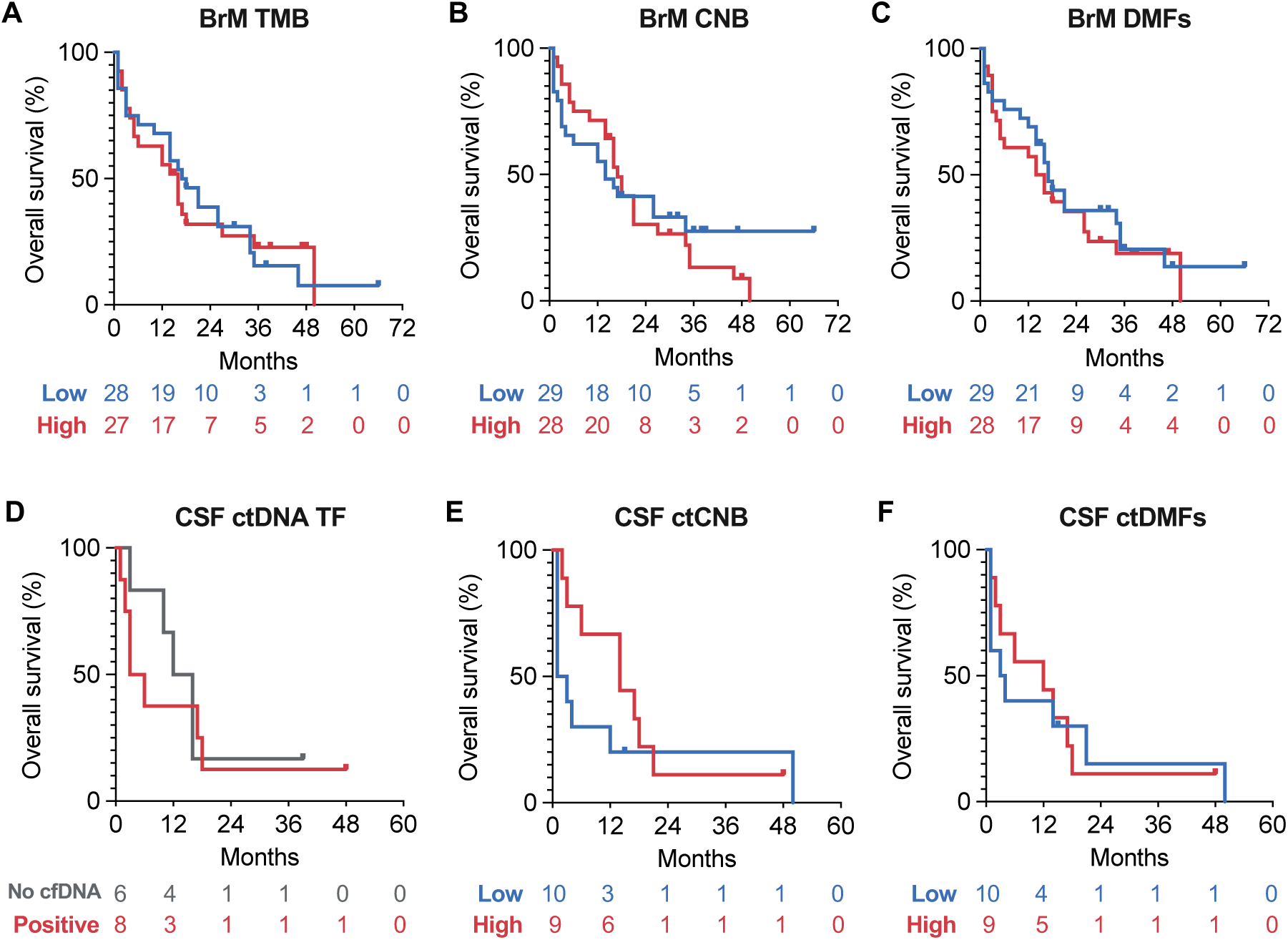
Overall survival based on molecular features of tissue and CSF. **(A-C)** Kaplan-Meier curves of patient OS stratified by TMB in (A), CNB in (B), and DMF counts in (C) detected in BrM tissues. **(D-F)** Kaplan-Meier curves of patient OS by CSF ctDNA-TF in (D), ctCNB in (E), and ctDMF counts in (F). All P values assessed by log-rank test are not significant (ns). Numbers at risk are displayed below the x-axis.

**Fig. S3.**
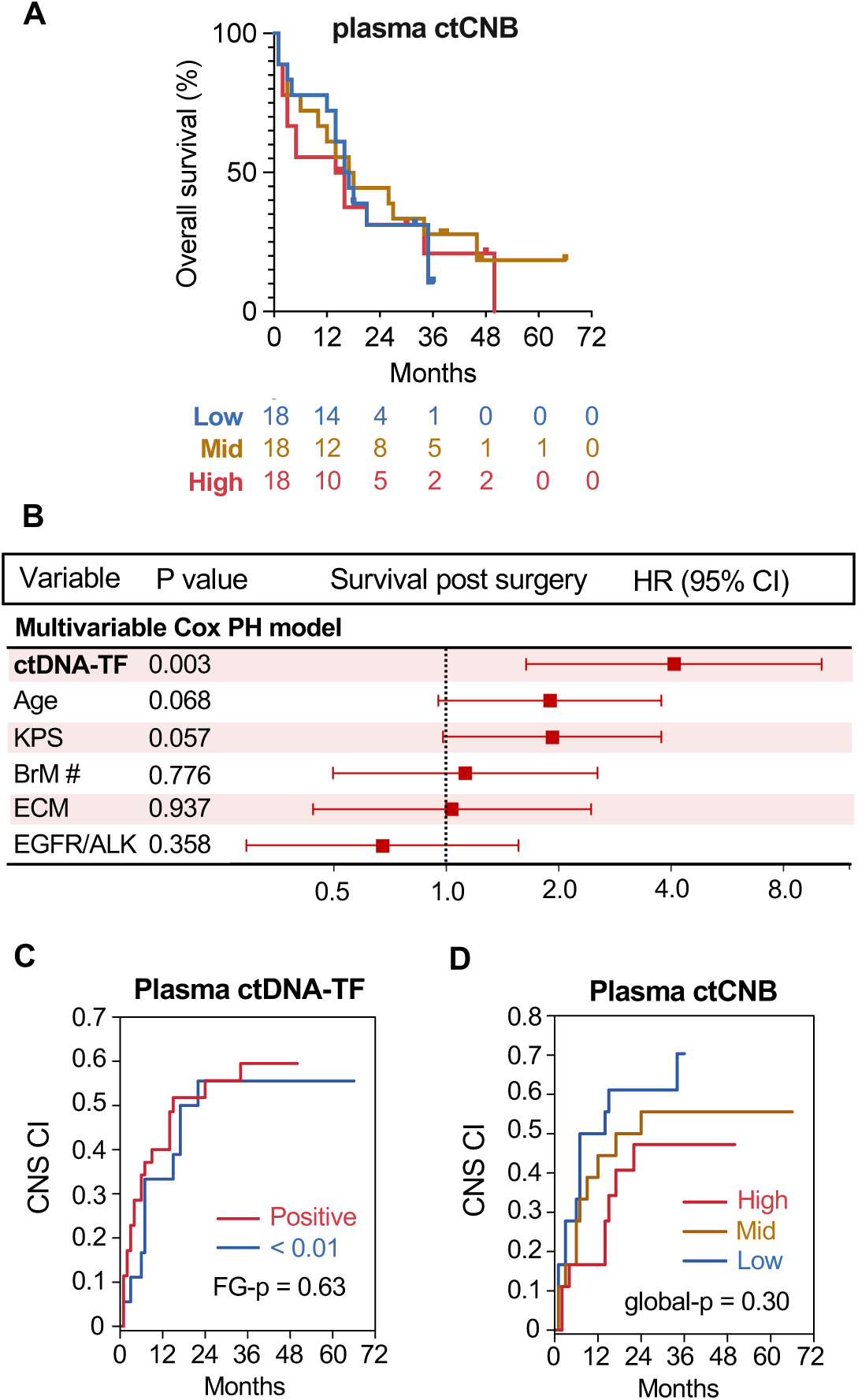
Plasma levels and patient outcomes. **(A)** Kaplan-Meier curves of patient OS by plasma ctCNB. P values assessed by log-rank test are not significant (ns). Numbers at risk are displayed below the x-axis. **(B)** Multivariate Cox proportional hazards regression of OS by plasma ctDNA-TF. HR, hazard ratio. CI, confidence interval. P values are calculated using a Wald test. **(C)** CIF of CNS progression stratified by plasma ctDNA-TF. P value calculated using a Fine-Gray model. N= 35 Positive, 19 <0.01. **(D)** CIF of CNS progression stratified by plasma ctCNB. N=18 High, 18 Mid, 18 Low. Overall P value calculated using omnibus Wald test for 3-group comparison (3-group-p).

**Fig. S4.**
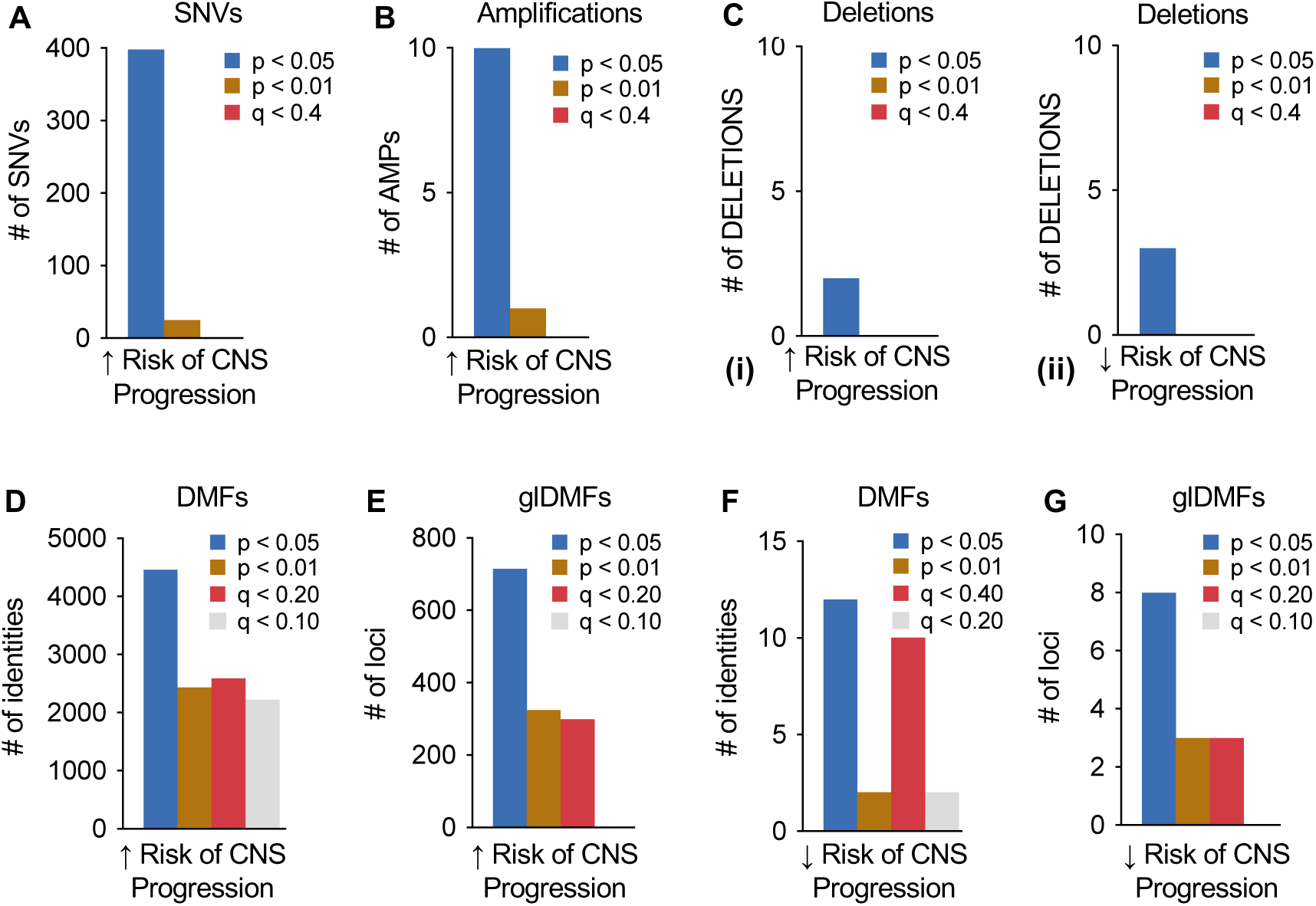
SNVs, CNVs, and DMFs associated with CNS progression. **(A)** Somatic variants (SNVs) in BrM tissue associated with increased patient risk of CNS progression by univariate Cox proportional hazards regression. **(B-C)** Gene-level copy number amplifications in (B) or deletions in (C) detected in BrM tissue associated with CNS progression risk by univariate Cox regression. Data in (A-D) based on variant calls and segment-level copy number changes on PredicineWES+ assay. **(D-E)** Plots of DMFs in (D) or gene-linked (gl-)DMFs in (E) detected in BrM tissue associated with increased risk of CNS progression by univariate Cox regression. **(F-G)** Plots of DMFs in (F) or gene-linked DMFs in (G) detected in BrM tissue associated with reduced risk of CNS progression by univariate Cox regression. Data in (D-G) based on fragment calls and gene annotations on PredicineEPIC assay. N=56 total cases (32 cases with subsequent CNS progression).

**Fig. S5.**
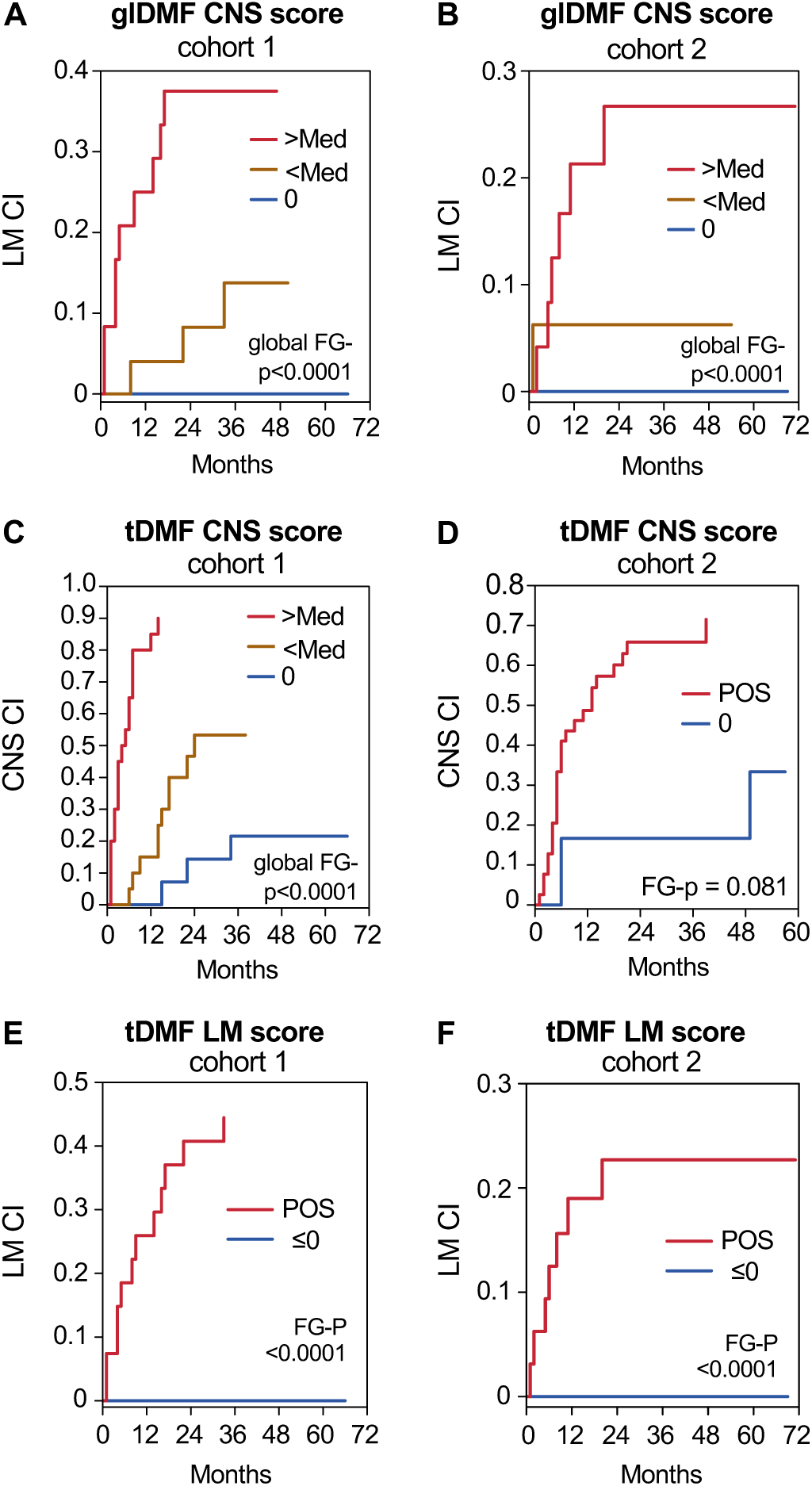
Extended analyses of methylation-based risk scores associated with CNS and LM progression. **(A)** Gene linked DMF CNS risk score was assessed for correlation with leptomeningeal metastasis (LM) specific progression. CIF of LM progression in cohort 1 patients was plotted based on CNS risk score. N=55 patients (24 High^+^ / 25 Low^+^ / 6=0). P value by Fine-Gray regression calculated using omnibus Wald test for global comparison (global FG-p<.0001). **(B)** CIF of LM progression in cohort 2 patients stratified by their CNS risk score (HR [Positive vs. 0] = not reportable, FG-p<0.0001). N= 45 total (24 High^+^/ 16 Low^+^ / 5=0). **(C)** All DMFs that correlated CNS progression were used to calculate a risk score (DMF CNS risk score) from BrM tissue. CIF of CNS progression in cohort 1 patients was plotted based on DMF CNS risk score (global FG-p=6.7x10^-6^). N=55 total (20 High^+^ / 20 Low^+^ / 5=0). **(D)** CIF of LM progression in cohort 2 stratified by their DMF CNS risk score (FG-p=0.081). N= 45 total (40 Positive / 5=0). **(E)** All DMFs correlated with LM were used to calculate a risk score (DMF LM risk score) from BrM tissue. CIF of LM specific progression in cohort 1 patients was plotted based on their DMF LM risk score (FG-p<0.0001). N=55 patients (27 High / 28 Low). **(F)** CIF of LM progression in cohort 2 stratified by DMF LM risk score threshold determined in (E) (FG-p<0.0001). N=45 total (32 High / 13 Low).

**Fig. S6.**
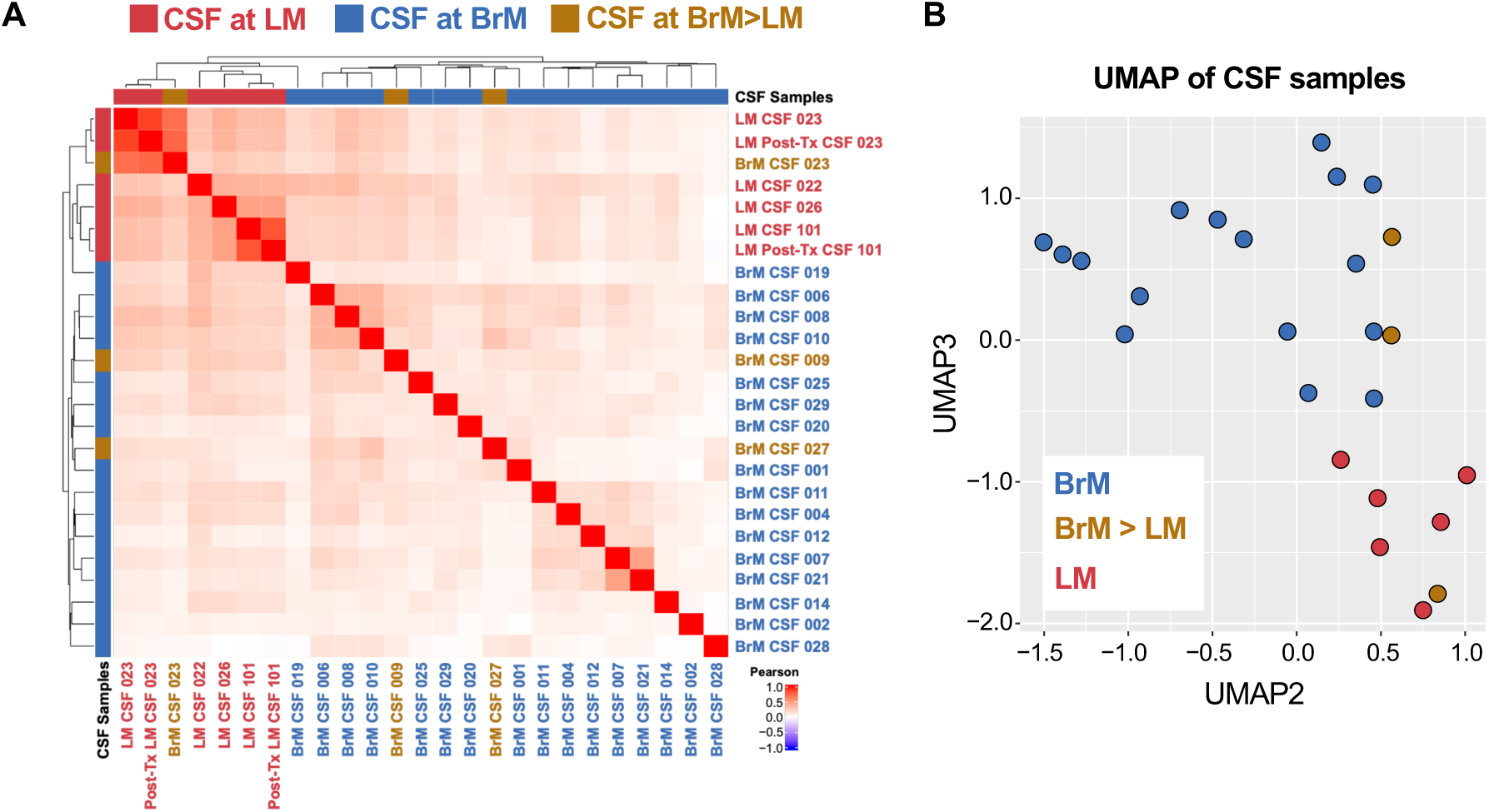
Comparison of CSF collected at time of brain parenchymal metastasis versus leptomeningeal metastasis. Heatmap **(A)** and UMAP **(B)** of CSF ctDNA methylation based on TCGA curated enriched tumor loci. CSF was collected at time of craniotomy or at LM diagnosis. BrM>NoLM: Patient with brain metastasis and no eventual LM progression, BrM>LM: Patient with brain metastasis and subsequent LM progression, LM: Patient with confirmed diagnosis of LM. Each point represents an individual sample.

**Fig. S7.**
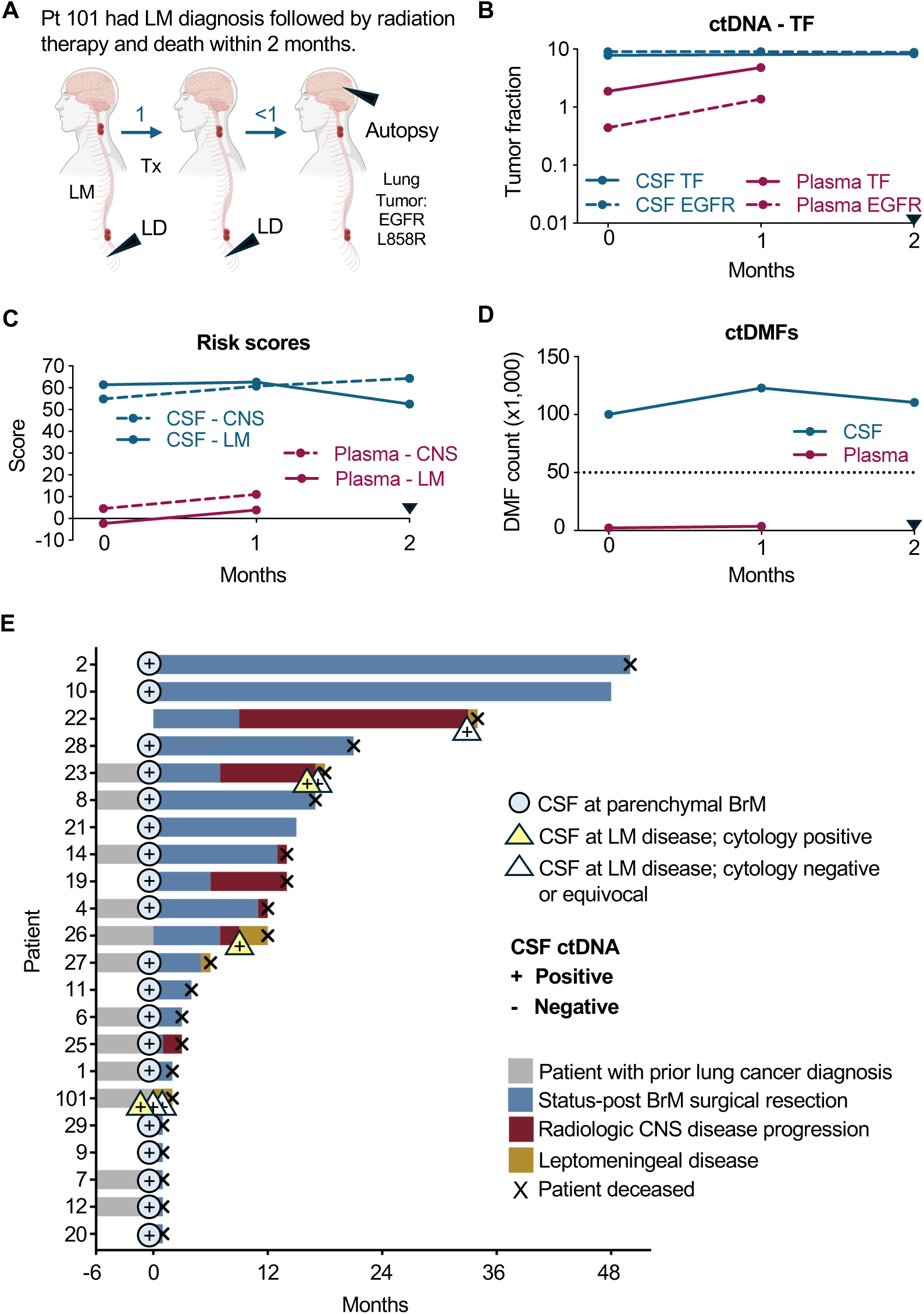
ctDNA features of CNS progression across longitudinal (liquid) CSF biopsies. **(A)** Overview of clinical course and sample collections from patient 32. Black arrows represent collections. LD, Lumbar drain. Tx, Treatment. N=1 Lung autopsy tissue, 2 PLA, and 3 CSFs samples in total. **(B-D)** ctDNA tumor fraction (TF) and EGFR L858R variant allele fraction (VAF) detected across CSF and plasma samples in (B), CNS and LM risk scores detected across CSF and plasma in (C), and total ctDMFs detected across CSF and plasma in (D) for patient 32. Black arrows represent time of the patient’s death. The dotted line in (D) represents DMF count of 50,000 fragments. **(E)** Swimmer plot illustrating the clinical course of CNS progression and CSF ctDNA assessments of patients with brain metastases and LM disease. N=22 patients.

## References and Notes

1. M. Valiente et al., The Evolving Landscape of Brain Metastasis. Trends Cancer 4, 176–196 (2018).

2. D. N. Cagney et al., Incidence and prognosis of patients with brain metastases at diagnosis of systemic malignancy: a population-based study. Neuro Oncol 19, 1511–1521 (2017).

3. L. E. Hendriks et al., EGFR mutated non-small cell lung cancer patients: more prone to development of bone and brain metastases? Lung Cancer 84, 86–91 (2014).

4. J. L. Villano et al., Incidence of brain metastasis at initial presentation of lung cancer. Neuro Oncol 17, 122–128 (2015).

5. Y. Kang, Y. Jin, Q. Li, X. Yuan, Advances in Lung Cancer Driver Genes Associated With Brain Metastasis. Front Oncol 10, 606300 (2020).

6. P. W. Sperduto et al., Survival in Patients With Brain Metastases: Summary Report on the Updated Diagnosis-Specific Graded Prognostic Assessment and Definition of the Eligibility Quotient. J Clin Oncol 38, 3773–3784 (2020).

7. N. Lamba, P. Y. Wen, A. A. Aizer, Epidemiology of brain metastases and leptomeningeal disease. Neuro Oncol 23, 1447–1456 (2021).

8. D. Li et al., Advances in targeted therapy in non-small cell lung cancer with actionable mutations and leptomeningeal metastasis. J Clin Pharm Ther 47, 24–32 (2022).

9. H. Cheng, R. Perez-Soler, Leptomeningeal metastases in non-small-cell lung cancer. Lancet Oncol 19, e43–e55 (2018).

10. Z. Pan et al., Leptomeningeal metastasis from solid tumors: clinical features and its diagnostic implication. Sci Rep 8, 10445 (2018).

11. P. K. Brastianos et al., Genomic Characterization of Brain Metastases Reveals Branched Evolution and Potential Therapeutic Targets. Cancer Discov 5, 1164–1177 (2015).

12. A. Skakodub et al., Genomic analysis and clinical correlations of non-small cell lung cancer brain metastasis. Nat Commun 14, 4980 (2023).

13. H. Gaitsch, R. J. M. Franklin, D. S. Reich, Cell-free DNA-based liquid biopsies in neurology. Brain 146, 1758–1774 (2023).

14. S. K. Cheok, et al., Tumor DNA Mutations From Intraparenchymal Brain Metastases Are Detectable in CSF. JCO Precis Oncol 5, (2021).

15. J. Wu et al., Cerebrospinal fluid circulating tumor DNA depicts profiling of brain metastasis in NSCLC. Mol Oncol 17, 810–824 (2023).

16. L. De Mattos-Arruda et al., Cerebrospinal fluid-derived circulating tumour DNA better represents the genomic alterations of brain tumours than plasma. Nat Commun 6, 8839 (2015).

17. J. A. Zuccato et al., Cerebrospinal fluid methylome-based liquid biopsies for accurate malignant brain neoplasm classification. Neuro Oncol 25, 1452–1460 (2023).

18. S. Klinsing et al., Detection of diagnostic somatic copy number alterations from cerebrospinal fluid cell-free DNA in brain tumor patients. Acta Neuropathol Commun 12, 177 (2024).

19. F. Iser et al., Cerebrospinal Fluid cfDNA Sequencing for Classification of Central Nervous System Glioma. Clin Cancer Res 30, 2974–2985 (2024).

20. M. M. Zheng et al., Cerebrospinal fluid circulating tumor DNA profiling for risk stratification and matched treatment of central nervous system metastases. Nat Med 31, 1547–1556 (2025).

21. A. P. Feinberg, B. Vogelstein, Hypomethylation distinguishes genes of some human cancers from their normal counterparts. Nature 301, 89–92 (1983).

22. D. Hu et al., Epigenetic Modifiers in Cancer Metastasis. Biomolecules 14, (2024).

23. A. Karlsson et al., Genome-wide DNA methylation analysis of lung carcinoma reveals one neuroendocrine and four adenocarcinoma epitypes associated with patient outcome. Clin Cancer Res 20, 6127–6140 (2014).

24. J. A. Karlow et al., Non-small Cell Lung Cancer Epigenomes Exhibit Altered DNA Methylation in Smokers and Never-smokers. Genomics Proteomics Bioinformatics 21, 991–1013 (2023).

25. F. Gimeno-Valiente et al., DNA methylation cooperates with genomic alterations during non-small cell lung cancer evolution. Nat Genet 57, 2226–2237 (2025).

26. J. A. Zuccato et al., Prediction of brain metastasis development with DNA methylation signatures. Nat Med 31, 116–125 (2025).

27. X. Yang et al., Gene body methylation can alter gene expression and is a therapeutic target in cancer. Cancer Cell 26, 577–590 (2014).

28. G. Lev Maor, A. Yearim, G. Ast, The alternative role of DNA methylation in splicing regulation. Trends Genet 31, 274–280 (2015).

29. J. R. M. Black et al., Longitudinal ultrasensitive ctDNA monitoring for high-resolution lung cancer risk prediction. Cell 188, 7083–7098 e7018 (2025).

30. C. Abbosh et al., Tracking early lung cancer metastatic dissemination in TRACERx using ctDNA. Nature 616, 553–562 (2023).

31. M. M. F. Schuurbiers et al., Recurrence prediction using circulating tumor DNA in patients with early-stage non-small cell lung cancer after treatment with curative intent: A retrospective validation study. PLoS Med 22, e1004574 (2025).

32. P. W. Sperduto et al., Estimating Survival in Patients With Lung Cancer and Brain Metastases: An Update of the Graded Prognostic Assessment for Lung Cancer Using Molecular Markers (Lung-molGPA). JAMA Oncol 3, 827–831 (2017).

33. D. Wasilewski et al., Clinical characteristics and outcomes in leptomeningeal disease with or without brain metastasis: insights from an explorative data analysis of the Charite LMD registry. J Neurooncol 175, 943–965 (2025).

34. E. L. Chang et al., Neurocognition in patients with brain metastases treated with radiosurgery or radiosurgery plus whole-brain irradiation: a randomised controlled trial. Lancet Oncol 10, 1037–1044 (2009).

35. P. D. Brown et al., Hippocampal Avoidance During Whole-Brain Radiotherapy Plus Memantine for Patients With Brain Metastases: Phase III Trial NRG Oncology CC001. J Clin Oncol 38, 1019–1029 (2020).

36. Q. T. Ostrom, C. H. Wright, J. S. Barnholtz-Sloan, Brain metastases: epidemiology. Handb Clin Neurol 149, 27–42 (2018).

37. N. Lamba et al., Racial disparities in supportive medication use among older patients with brain metastases: a population-based analysis. Neuro Oncol 22, 1339–1347 (2020).

38. J. A. Wilcox et al., Leptomeningeal metastases from solid tumors: A Society for Neuro-Oncology and American Society of Clinical Oncology consensus review on clinical management and future directions. Neuro Oncol 26, 1781–1804 (2024).

39. M. Vaz Batista et al., The DEBBRAH trial: Trastuzumab deruxtecan in HER2-positive and HER2-low breast cancer patients with leptomeningeal carcinomatosis. Med 6, 100502 (2025).

40. T. J. Yang et al., Clinical trial of proton craniospinal irradiation for leptomeningeal metastases. Neuro Oncol 23, 134–143 (2021).

41. M. D. White et al., Detection of Leptomeningeal Disease Using Cell-Free DNA From Cerebrospinal Fluid. JAMA Netw Open 4, e2120040 (2021).

42. K. Bai et al., Cerebrospinal fluid circulating tumour DNA genotyping and survival analysis in lung adenocarcinoma with leptomeningeal metastases. J Neurooncol 165, 149–160 (2023).

43. T. D. Azad et al., Quantification of cerebrospinal fluid tumor DNA in lung cancer patients with suspected leptomeningeal carcinomatosis. NPJ Precis Oncol 8, 121 (2024).

44. T. A. Bale et al., Clinical Experience of Cerebrospinal Fluid-Based Liquid Biopsy Demonstrates Superiority of Cell-Free DNA over Cell Pellet Genomic DNA for Molecular Profiling. J Mol Diagn 23, 742–752 (2021).

45. A. M. Omuro et al., High incidence of disease recurrence in the brain and leptomeninges in patients with nonsmall cell lung carcinoma after response to gefitinib. Cancer 103, 2344–2348 (2005).

46. Y. S. Li et al., Leptomeningeal Metastases in Patients with NSCLC with EGFR Mutations. J Thorac Oncol 11, 1962–1969 (2016).

47. C. Riviere-Cazaux et al., Longitudinal Glioma Monitoring via Cerebrospinal Fluid Cell-Free DNA. Clin Cancer Res 31, 881–889 (2025).

48. Y. Choi et al., Circulating Tumor DNA Dynamics Reveal KRAS G12C Mutation Heterogeneity and Response to Treatment with the KRAS G12C Inhibitor Divarasib in Solid Tumors. Clin Cancer Res 30, 3788–3797 (2024).

49. A. A. Davis et al., Genomic Complexity Predicts Resistance to Endocrine Therapy and CDK4/6 Inhibition in Hormone Receptor-Positive (HR+)/HER2-Negative Metastatic Breast Cancer. Clin Cancer Res 29, 1719–1729 (2023).

